# Networks of information token recurrences derived from genomic sequences may reveal hidden patterns in epidemic outbreaks: A case study of the 2019-nCoV coronavirus

**DOI:** 10.1101/2020.02.07.20021139

**Authors:** Markus Luczak-Roesch

## Abstract

Profiling the genetic evolution and dynamic spreading of viruses is a crucial task when responding to epidemic outbreaks. We aim to devise novel ways to model, visualise and analyse the temporal dynamics of epidemic outbreaks in order to help researchers and other people involved in crisis response to make well-informed and targeted decisions about from which geographical locations and time periods more genetic samples may be required to fully understand the outbreak. Our approach relies on the application of Transcendental Information Cascades to a set of temporally ordered nucleotide sequences, and we apply it to real-world data that was collected during the currently ongoing outbreak of the novel 2019-nCoV coronavirus. We assess information-theoretic and network-theoretic measures that characterise the resulting complex network and identify touching points and temporal pathways that are candidates for deeper investigation by geneticists and epidemiologists.

## Introduction

How far a virus will eventually spread at local, regional, national and international levels is of central concern during an epidemic outbreak due to the severe consequences large-scale epidemics have on human well-being [5], the stability and coherence of social systems [11], and the global economy [23]. A thorough understanding of the genetic characteristics of a virus is crucial to understand the way it is (or may be) transmitted (e.g. human-to-human transmissibility) in order to be able to anticipate the potential reach of an outbreak and develop effective countermeasures. The recent outbreak of a new type of coronavirus – 2019-nCoV – provides plenty of examples of the way researchers seek to quickly approach the aforementioned problems of genetic decomposition of the virus [2, 8] and modelling of its transmission dynamics [15].

Here we present a novel approach to gain insights into the transmission dynamics of an epidemic outbreak. Our method is based on Transcendental Information Cascades [16,17] and combines gene sequence analysis with temporal data mining to uncover potential relationships between a virus’ genetic evolution and distinct occurrences of infections. We apply this new approach to publicly available genomic sequence data from the currently ongoing outbreak of the 2019-nCoV coronavirus.

Our results show pathways of similarity between certain clusters of 2019-nCoV virus samples taken in different geographical locations, and indicate coronavirus cases that are candidates for further investigation into the human touch points of these patients in order to derive a detailed understanding of how the virus may have spread, and how and why it has genetically evolved. Our analysis provides a unique view to the dynamics of the 2019-nCoV coronavirus outbreak that complements knowledge obtained using other state-of-the-art methods such as Bayesian evolutionary analysis [9].

## Materials and methods

### Research data and preprocessing

We obtained a total of 87 genomic sequences from human subjects that were available via the *GISAID EpiFlu Database*™ ^1^ as of Monday, February 10th 2020.

These data were preprocessed using the R programming language. First, we derived a metadata object that allows to store the sequence identifier, the collection location, the collection date, and the raw nucleotide sequence. The metadata object was then filtered to keep only those sequences for which the collection date was not empty and which featured a length that fell within a margin of 1, 000 of the estimated 30, 000 nucleotides that are currently assumed to make up the genomic code of the 2019-nCoV coronavirus (i.e. we cover the range of nucleotide sequences from 29, 000 to 31, 000 in length). This left us with a total of 82 genomic sequences. We then exported a single CSV file containing only the raw nucleotide sequences in ascending order by the collection date of the sample. This structure is the standard input format for the *genes-CODON-samplesequence* tokeniser featured in the Transcendental Information Cascades R toolchain that is available as free and open scientific software^2^.

### Construction of Transcendental Information Cascades

Transcendental Information Cascades (TICs) are a method that first transforms any kind of sequential data into a network of recurring *information tokens*, to then exploit this network as a kind of analytical middle ground between views to the source data that would otherwise only be accessible individually through distinct analytical frameworks [16, 17]. The approach falls into the category of contemporary approaches that make use of network theory for the study of dynamical complex systems [4, 19, 21]. In the field of phylogenetic and phylogeographic analysis our approach can be compared to haplotype networks for example [1, 18] but with the benefit that the resulting network preserves access to temporal patterns at both macroscopic (e.g. whole generation time periods) and microscopic (e.g. short time period bursts) resolution.

At an abstract level a Transcendental Information Cascade is constructed from some sequentially ordered source data in the following way: At first the data from each distinct sequence step is processed to extract unique information tokens. These tokens form the *identifier set* for that particular sequence step. Then a directed network is generated with one vertex for each sequence step and edges between any two vertices that have a particular token from their identifier set in common and that does not occur in any identifier set of vertices that represent sequence steps between these two. In other words, identifier sets represent information token co-occurrences and edges represent information token recurrences.

As the information tokens of interest in this study we encode unique codon identifiers as (a) their position when sliding a window of size 3 in steps of size 3 over the nucleotide sequence (e.g. “pos1”, “pos2”, …), (b) a flag that indicates the reading frame (“+1”, “+2” or “+3”) that captures the respective triplet in the current window, and (c) the actual matched nucleotide triplet that constitutes the matched codon. The use of this tokenisation is motivated by previous work on phylogenetic profiling and gene sequencing [6, 13] that suggested that codons suit well for assessing similarities (and dissimilarities) at the gene, chromosome, or genome levels [6, 14, 22].

An example of the particular encoding we use is visualised in Figure 1. It is this tokenisation of the original nucleotide sequences that we use in the TIC approach to construct the information token recurrence network from. This means in our TIC instantiation the nodes represent distinct nucleotide sequences of the virus obtained from patient samples ordered by the date of their collection by researchers (random order in case of same date collections). The edges are unique codon identifier recurrences matched between different nucleotide sequences.

**Figure 1.**
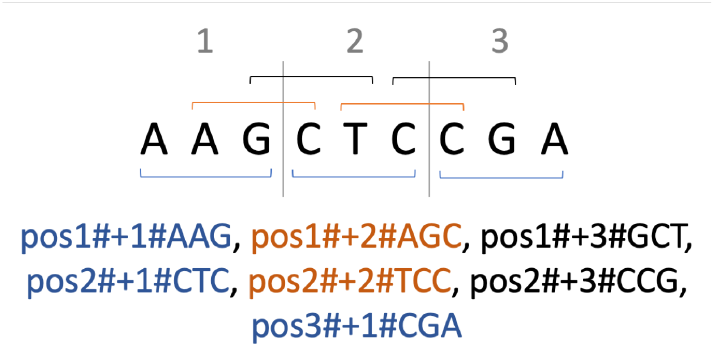
Example of the unique codon tokenisation used in this study. The example sequence comprises three window steps and shows how the +1 (marked in blue), +2 (marked in orange) and +3 (marked in black) reading frames lead to seven unique codon tokens extracted from the nucleotide sequence.

### Analysis of Transcendental Information Cascades

The TIC network we derive in the way described before can be analysed in a variety of ways. Information-theoretic and network-theoretic measures have been introduced as the base framework to start the analysis from [16]. In particular we perform (1) an analysis of the information token entropy and evenness over time, (2) an analysis of network clusters of sequences that can be detected in the TIC network, and (3) an analysis of the intra-cluster and inter-cluster nucleotide similarity of the original sequences.

#### Information token entropy and evenness

We perform an analysis of information token entropy and information token evenness in order to understand whether the virus evolution can be considered an open or a closed system. Therefore we assess both measures in an accumulated fashion for each progression step through the ordered nucleotide sequences. In a closed system the information token entropy should strive towards an equilibrium of maximum entropy and not feature any wave-like up and down patterns. If we observe wave-like patterns it means nucleotide sequences at later progression stages feature codon identifiers that have not been observed in earlier nucleotide sequences.

So to measure information token entropy we assess which unique codon identifiers were seen up to a progression step and how often these were seen. This allows to compute the probability for every codon identifier, which then can be used to determine the information token entropy up to that point following the definition of Shannon entropy given as follows:

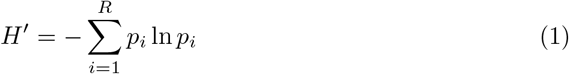

Analog to this we can then assess the information token evenness as per Pielou’s species evenness index, which is defined as

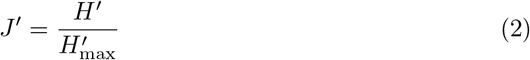

where *H*^*’*^ is the measured Shannon entropy, and 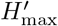 is the maximum entropy that would be measured if all tokens would be equally likely to occur.

#### Network analysis

The Transcendental Information Cascade network we constructed can be mathematically and visually analysed in order to find structures that are of particular significance for a single epidemic outbreak or to compare different outbreaks structurally. In this study we focus on the former due to the lack of comparative data about other epidemic outbreaks.

We first perform a cluster analysis using the random walk apporach by Blondel et al. [3] on the weighted network that we can construct from our original TIC network by collapsing all edges between the same vertices to unique edges weighted by the sum of the collapsed edges (see Figure 2 for a schematic example and Figure 4 for the actual networks we construct following this approach). Afterwards, we analyse visually the network using the open source software Gephi. In Gephi we scale vertices by their degree and edges by their weight. Furthermore, we colour vertices by their cluster membership. We then run the *Yifan Hu* [12] layout algorithm (optimal distance: 10,000; relative strength: 0.2; initial step size: 20; step ratio: 0.95; quadtree max level: 50; theta: 0.8; convergence threshold: 1*10^*−*4^; adaptive cooling: enabled) to visualise the network. We repeat the exact same process to construct and visualise three further networks (cf. Figures 5-7) that are representative of the TIC when exclusively focusing on the codon identifiers of each of the reading frames individually.

**Figure 2.**
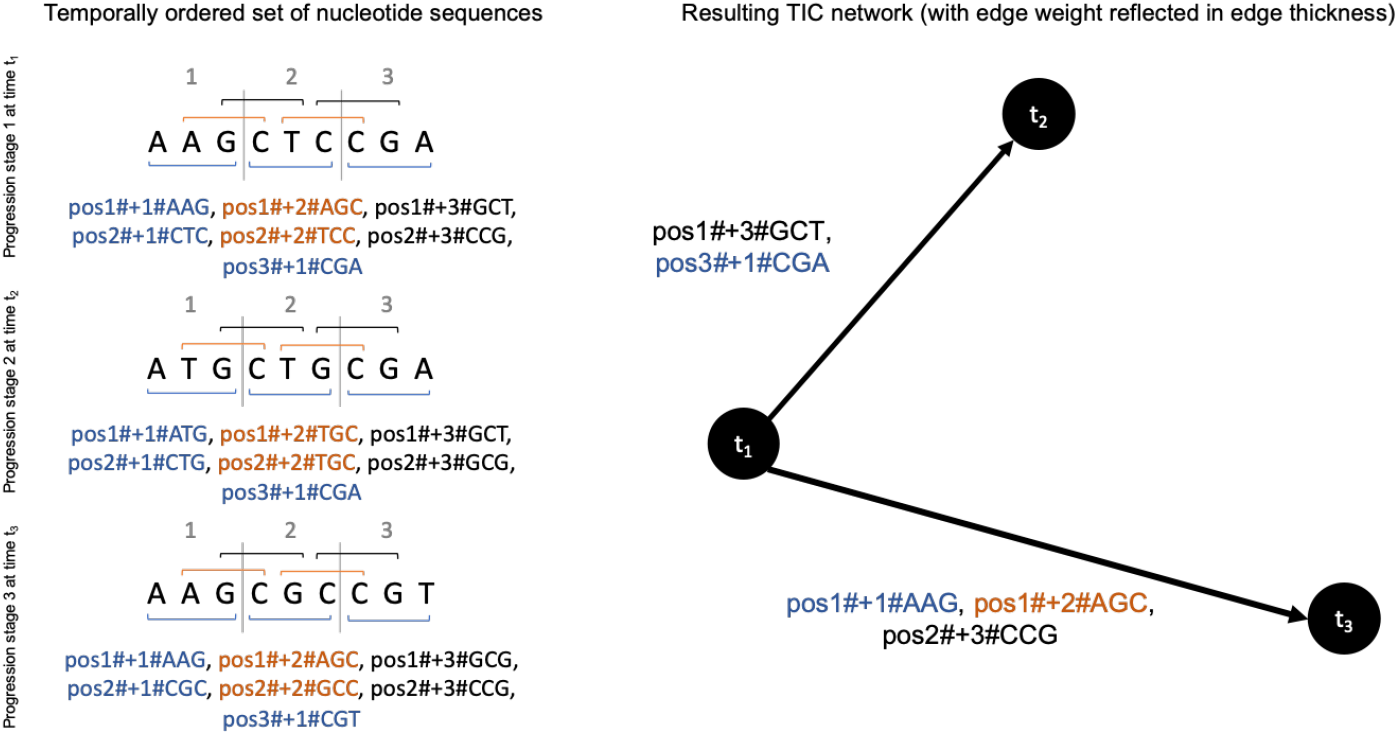
Schematic example of the weighted TIC network constructed from nucleotide sequences with tokenised unique codon identifiers.

**Figure 3.**
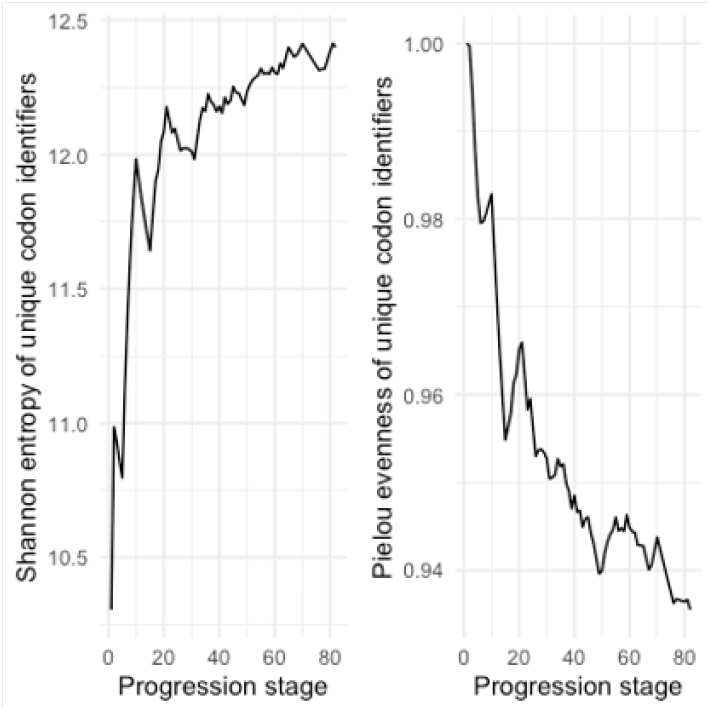
Information-theoretic assessment. Both entropy and evenness graphs feature wave-like patterns, indicating that the set of observed unique codon identifiers grows with increasing progression stages while the distribution of all observed unique codon identifiers remains rather even.

**Figure 4.**
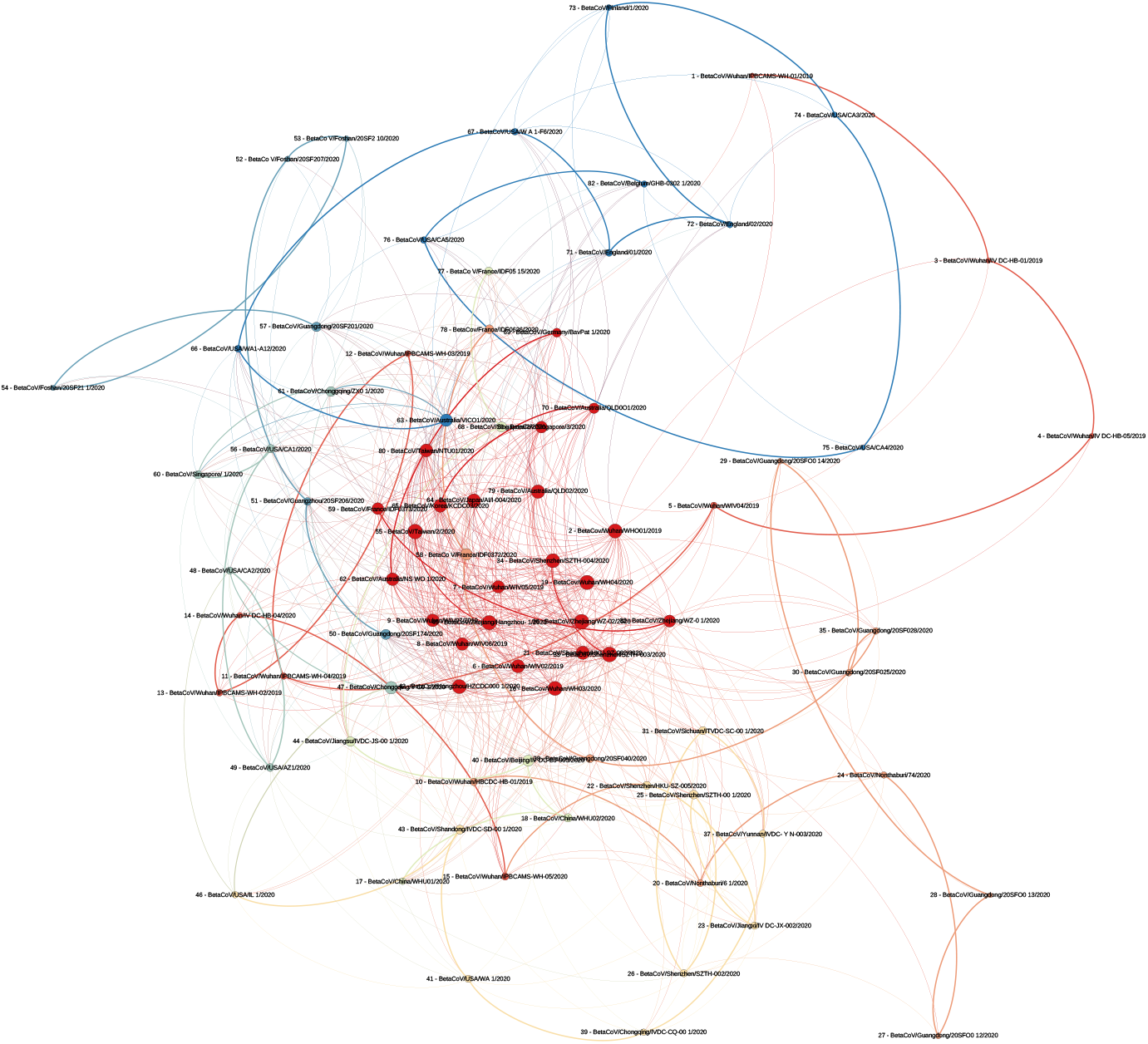
Figure 4. Weighted TIC network. From the original TIC network we generate a weighted network by collapsing all edges between the same vertices to unique edges weighted by the sum of the collapsed edges. The colours represent the nine identified network clusters. Node size represents the degree and edge thickness represents edge weight (in the sense that higher edge weight indicates a higher amount of codon identifiers shared by two vertices). Numeric indices before the genome identifiers represent the progression stage index of the particular vertex in the sequence of all ordered nucleotide sequences we analysed.

**Figure 5.**
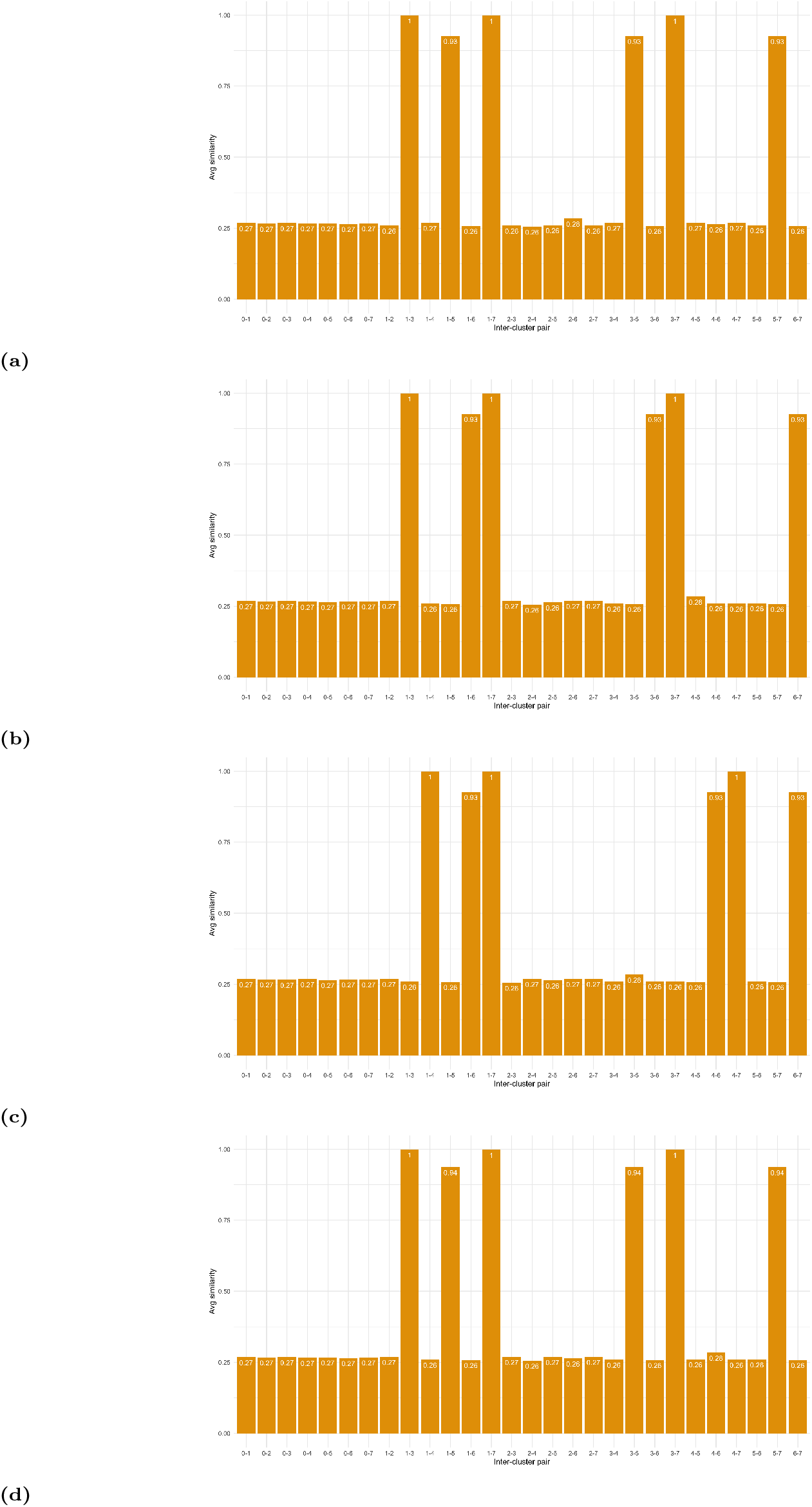
Overview of inter-cluster similarities for (a) the full TIC network, (b) the +1 reading frame network, (c) the +2 reading frame network, and (d) the +3 reading frame network.

**Figure 6.**
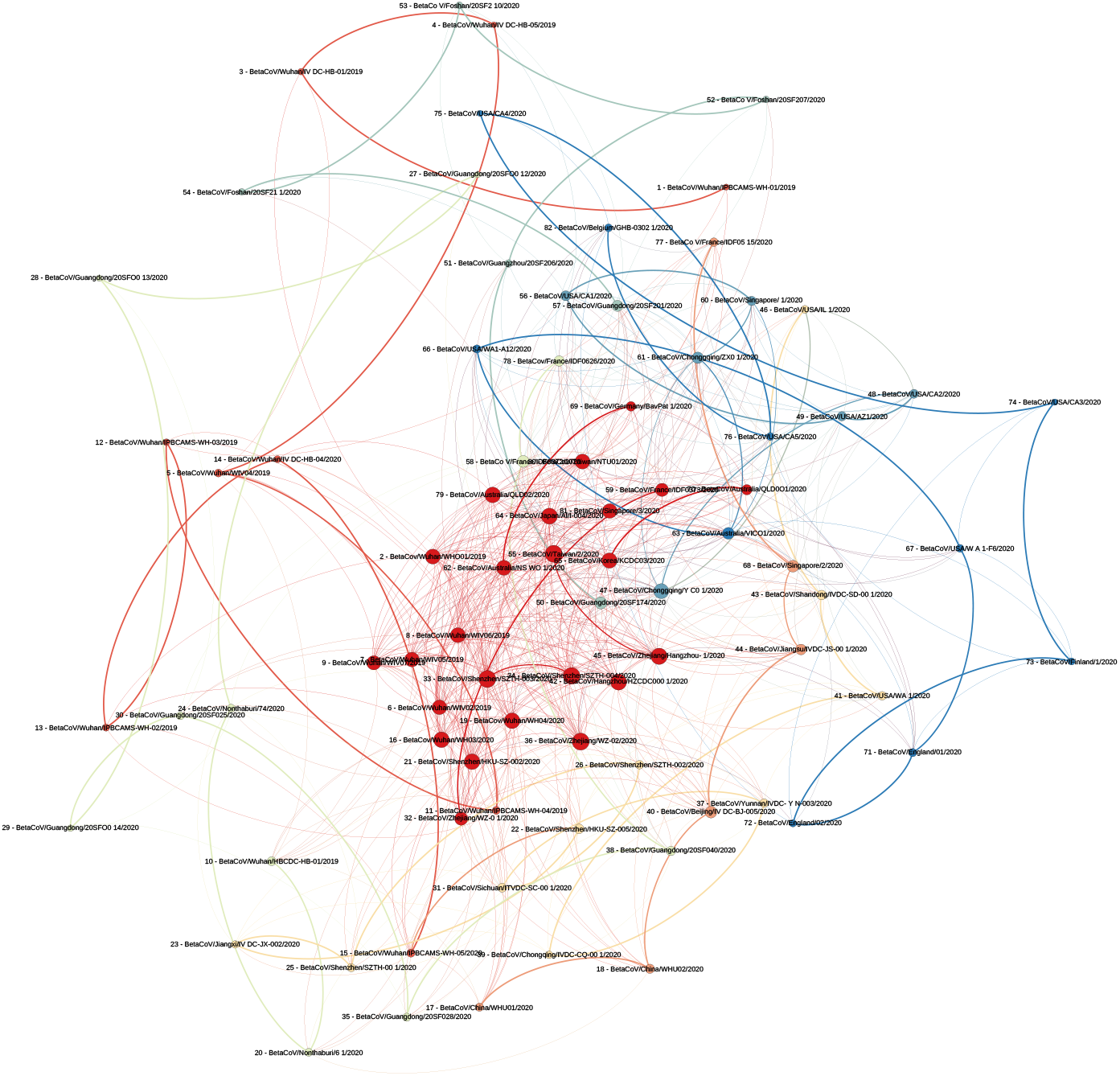
+1 reading frame network.

**Figure 7.**
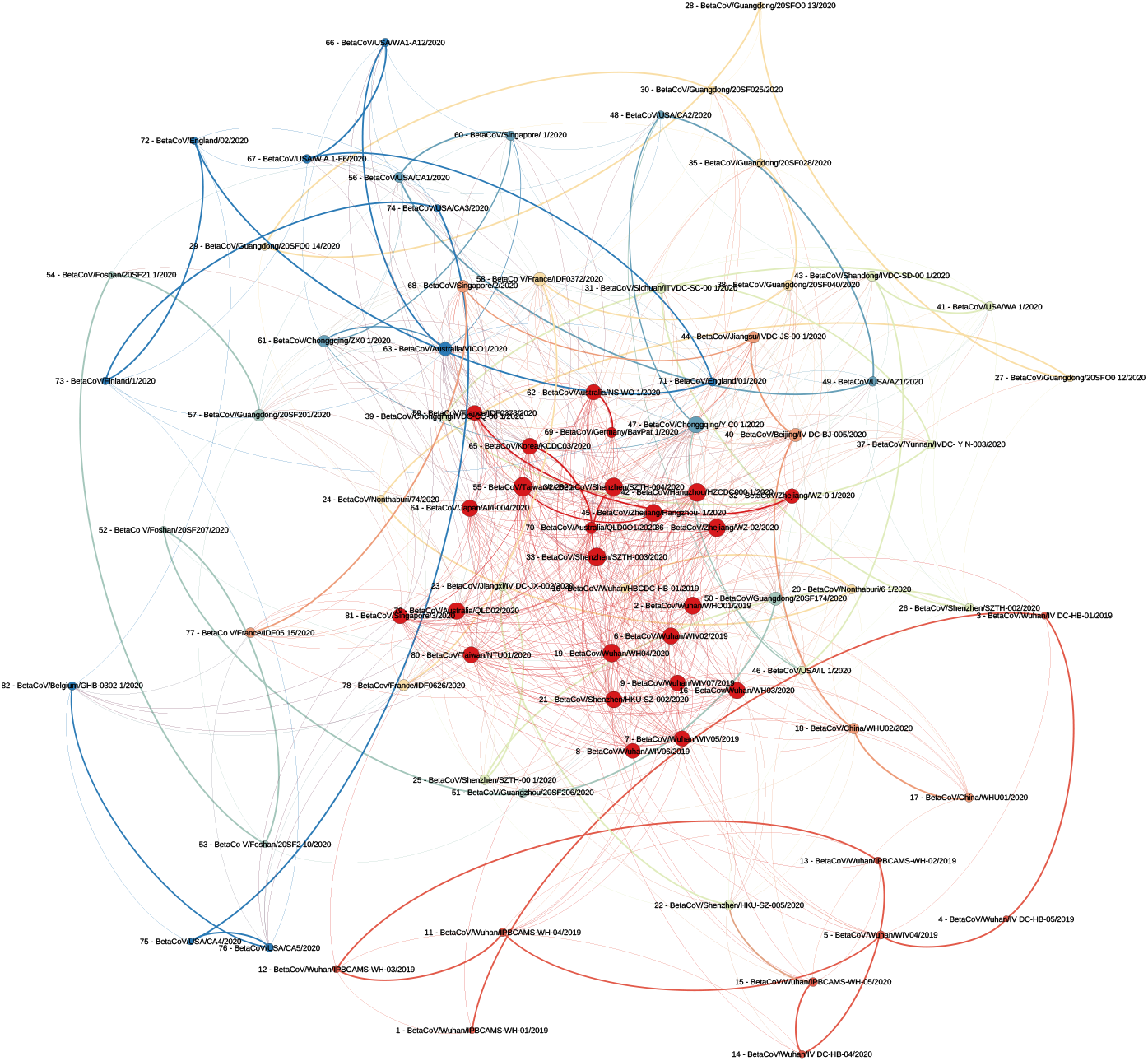
+2 reading frame network.

#### Cluster evaluation

In order to evaluate the meaningfulness of the TIC network against some baseline, we computer the intra-cluster and inter-cluster similarity of the raw nucleotide sequences. This is done by computing all pairwise comparisons of sequences within individual clusters (intra-cluster similarity) and comparison between all sequences from a cluster with all sequences that are not within that cluster (inter-cluster similarity) using the *compareStrings* function provided by the Biostrings R package [20]. The similarity of two sequences is then simply defined as follows:

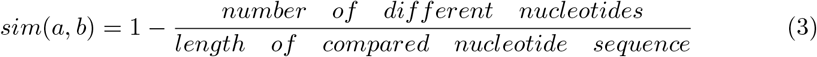

It is important to note that not all sequences we obtained had the exact same length (mean length of nucleotide sequences: 29861.95; standard deviation: 28.40705). Hence, we performed this comparison only on the sub-sequence from the first nucleotide position to the maximum nucleotide position of the shorter of the two sequences *a* and *b*.

## Results and discussion

### Information-theoretic analysis

Figure 3 shows the graphs for the information token entropy and information token evenness. As one can see, these feature the aforementioned wave-like patterns, which means the system can be regarded *open* in the sense that over time the number of unique codon identifiers grows.

It is important to note that the evenness of the distribution of codon identifiers is relatively high, converging around a value of 0.95. This means that those codon identifiers that have been observed are quite evenly distributed.

These results allow the interpretation that there is evolving variation in the genomic sequences we studied, but (b) there is no indication for the dominance of a particular variance.

### Network analysis

The weighted network derived from the original TIC network features a modularity of 0.762 and a diameter of 7, which mainly reflects its small size and its particular structure that results from preserving temporal order in the sense that edges can only ever direct forward in time (i.e. to vertices that represent a later stage). However, with an average degree of 8.671 we find that there is certainly structure that differs from just simple step-by-step forward paths that lead to the consecutively next node. This latter result suggests that some codon paths branch and merge, emphasising that there is an evolving variation in the studied genomic sequences.

From the cluster analysis in combination with a visual inspection of the resulting graph (please refer to Figure 6 as well as Table 11 in the appendix) we find the following patterns that are noteworthy:

**Table 1.** Intra-cluster similarities.

| Cluster | Avg. similarity | SD | Min. similarity | Max. similarity | Cluster size |
| --- | --- | --- | --- | --- | --- |
| 0 | 0.277353 | 0.088247 | 0.250620 | 0.999866 | 24 |
| 1 | 0.999730 | 0.000222 | 0.999231 | 1.000000 | 9 |
| 2 | 0.999915 | 0.000048 | 0.999832 | 1.000000 | 11 |
| 3 | 0.999385 | 0.000371 | 0.998461 | 0.999933 | 10 |
| 4 | 0.998768 | 0.001345 | 0.996387 | 1.000000 | 6 |
| 5 | 0.883701 | 0.112398 | 0.781976 | 0.999899 | 6 |
| 6 | 0.999926 | 0.000036 | 0.999866 | 1.000000 | 6 |
| 7 | 0.997900 | 0.002419 | 0.991158 | 1.000000 | 10 |

**Table 2.**
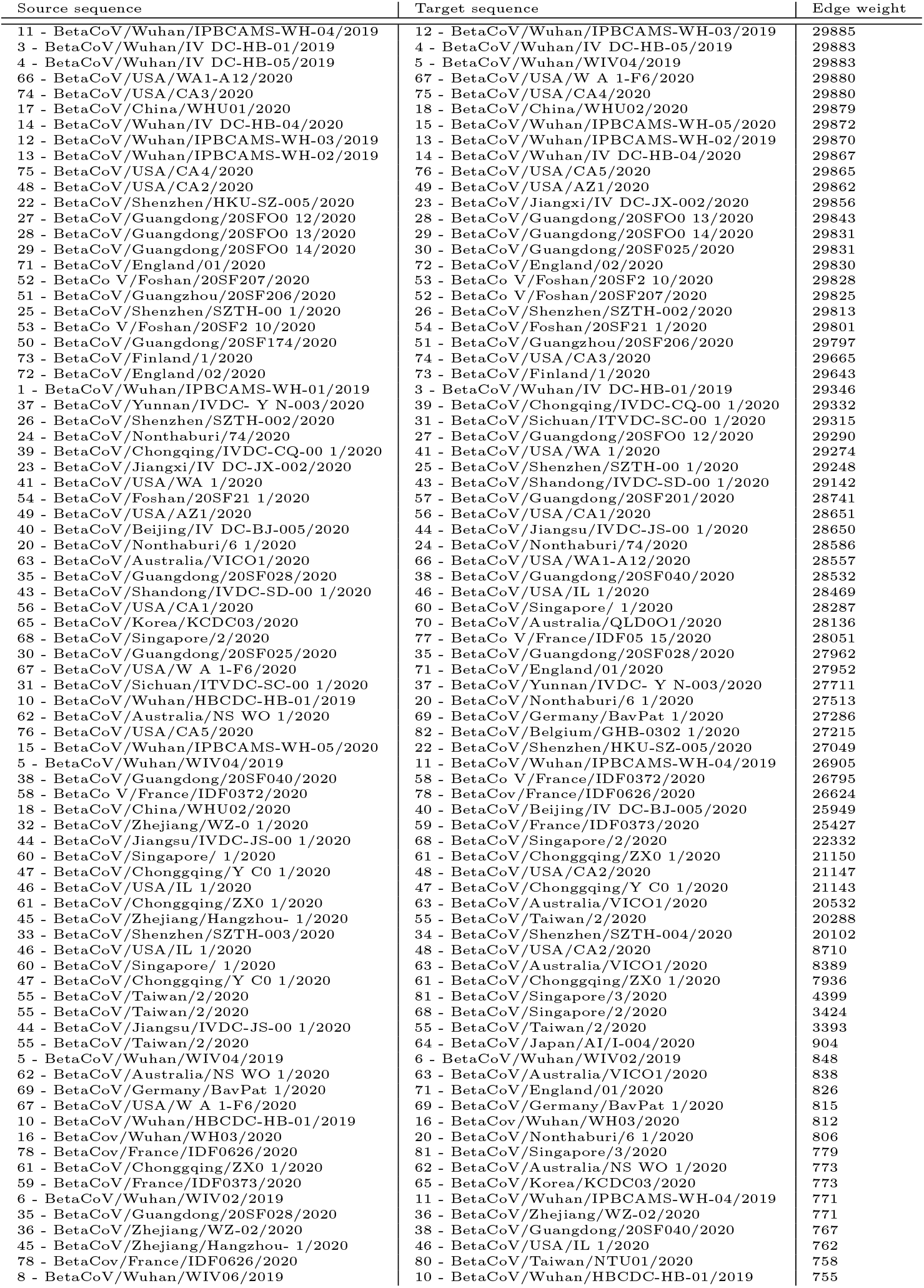
Edge weight between distinct nucleotide sequences in the Transcendental Information Cascade we constructed.

**Table 3.** Edge weight between distinct nucleotide sequences in the Transcendental Information Cascade we constructed.

| Source sequence | Target sequence | Edge weight |
| --- | --- | --- |
| 43 - BetaCoV/Shandong/IVDC-SD-00 1/2020 | 45 - BetaCoV/Zhejiang/Hangzhou- 1/2020 | 754 |
| 19 - BetaCov/Wuhan/WH04/2020 | 22 - BetaCoV/Shenzhen/HKU-SZ-005/2020 | 748 |
| 34 - BetaCoV/Shenzhen/SZTH-004/2020 | 36 - BetaCoV/Zhejiang/WZ-02/2020 | 747 |
| 57 - BetaCoV/Guangdong/20SF201/2020 | 58 - BetaCoV/France/IDF0372/2020 | 742 |
| 69 - BetaCoV/Germany/BavPat 1/2020 | 79 - BetaCoV/Australia/QLD02/2020 | 740 |
| 15 - BetaCoV/Wuhan/IPBCAMS-WH-05/2020 | 19 - BetaCov/Wuhan/WH04/2020 | 731 |
| 20 - BetaCoV/Nonthaburi/6 1/2020 | 21 - BetaCoV/Shenzhen/HKU-SZ-002/2020 | 727 |
| 6 - BetaCoV/Wuhan/WIV02/2019 | 7 - BetaCoV/Wuhan/WIV05/2019 | 725 |
| 21 - BetaCoV/Shenzhen/HKU-SZ-002/2020 | 24 - BetaCoV/Nonthaburi/74/2020 | 715 |
| 80 - BetaCoV/Taiwan/NTU01/2020 | 81 - BetaCoV/Singapore/3/2020 | 714 |
| 9 - BetaCoV/Wuhan/WIV07/2019 | 17 - BetaCoV/China/WHU01/2020 | 713 |
| 5 - BetaCoV/Wuhan/WIV04/2019 | 7 - BetaCoV/Wuhan/WIV05/2019 | 711 |
| 7 - BetaCoV/Wuhan/WIV05/2019 | 19 - BetaCov/Wuhan/WH04/2020 | 703 |
| 76 - BetaCoV/USA/CA5/2020 | 79 - BetaCoV/Australia/QLD02/2020 | 700 |
| 79 - BetaCoV/Australia/QLD02/2020 | 82 - BetaCoV/Belgium/GHB-0302 1/2020 | 700 |
| 7 - BetaCoV/Wuhan/WIV05/2019 | 11 - BetaCoV/Wuhan/IPBCAMS-WH-04/2019 | 698 |
| 41 - BetaCoV/USA/WA 1/2020 | 42 - BetaCoV/Hangzhou/HZCDC000 1/2020 | 698 |
| 42 - BetaCoV/Hangzhou/HZCDC000 1/2020 | 43 - BetaCoV/Shandong/IVDC-SD-00 1/2020 | 698 |
| 63 - BetaCoV/Australia/VICO1/2020 | 64 - BetaCoV/Japan/AI/1-004/2020 | 694 |
| 38 - BetaCoV/Guangdong/20SF040/2020 | 50 - BetaCoV/Guangdong/20SF174/2020 | 693 |
| 68 - BetaCoV/Singapore/2/2020 | 70 - BetaCoV/Australia/QLD001/2020 | 687 |
| 2 - BetaCov/Wuhan/WHO01/2019 | 9 - BetaCoV/Wuhan/WIV07/2019 | 686 |
| 21 - BetaCoV/Shenzhen/HKU-SZ-002/2020 | 33 - BetaCoV/Shenzhen/SZTH-003/2020 | 685 |
| 21 - BetaCoV/Shenzhen/HKU-SZ-002/2020 | 50 - BetaCoV/Guangdong/20SF174/2020 | 685 |
| 55 - BetaCoV/Taiwan/2/2020 | 56 - BetaCoV/USA/CA1/2020 | 685 |
| 36 - BetaCoV/Zhejiang/WZ-02/2020 | 50 - BetaCoV/Guangdong/20SF174/2020 | 678 |
| 55 - BetaCoV/Taiwan/2/2020 | 62 - BetaCoV/Australia/NS WO 1/2020 | 677 |
| 49 - BetaCoV/USA/AZ1/2020 | 55 - BetaCoV/Taiwan/2/2020 | 676 |
| 65 - BetaCoV/Korea/KCDC03/2020 | 68 - BetaCoV/Singapore/2/2020 | 675 |
| 70 - BetaCoV/Australia/QLD001/2020 | 77 - BetaCoV/France/IDF05 15/2020 | 675 |
| 16 - BetaCov/Wuhan/WH03/2020 | 21 - BetaCoV/Shenzhen/HKU-SZ-002/2020 | 674 |
| 64 - BetaCoV/Japan/AI/1-004/2020 | 66 - BetaCoV/USA/WA1-A12/2020 | 674 |
| 62 - BetaCoV/Australia/NS WO 1/2020 | 64 - BetaCoV/Japan/AI/1-004/2020 | 671 |
| 19 - BetaCov/Wuhan/WH04/2020 | 42 - BetaCoV/Hangzhou/HZCDC000 1/2020 | 666 |
| 42 - BetaCoV/Hangzhou/HZCDC000 1/2020 | 45 - BetaCoV/Zhejiang/Hangzhou- 1/2020 | 665 |
| 8 - BetaCoV/Wuhan/WIV06/2019 | 16 - BetaCov/Wuhan/WH03/2020 | 664 |
| 21 - BetaCoV/Shenzhen/HKU-SZ-002/2020 | 36 - BetaCoV/Zhejiang/WZ-02/2020 | 654 |
| 42 - BetaCoV/Hangzhou/HZCDC000 1/2020 | 64 - BetaCoV/Japan/AI/1-004/2020 | 654 |
| 9 - BetaCoV/Wuhan/WIV07/2019 | 32 - BetaCoV/Zhejiang/WZ-0 1/2020 | 651 |
| 30 - BetaCoV/Guangdong/20SF025/2020 | 33 - BetaCoV/Shenzhen/SZTH-003/2020 | 646 |
| 18 - BetaCoV/China/WHU02/2020 | 32 - BetaCoV/Zhejiang/WZ-0 1/2020 | 641 |
| 64 - BetaCoV/Japan/AI/1-004/2020 | 69 - BetaCoV/Germany/BavPat 1/2020 | 637 |
| 42 - BetaCoV/Hangzhou/HZCDC000 1/2020 | 62 - BetaCoV/Australia/NS WO 1/2020 | 630 |
| 32 - BetaCoV/Zhejiang/WZ-0 1/2020 | 40 - BetaCoV/Beijing/IV DC-BJ-005/2020 | 629 |
| 34 - BetaCoV/Shenzhen/SZTH-004/2020 | 35 - BetaCoV/Guangdong/20SF028/2020 | 628 |
| 7 - BetaCoV/Wuhan/WIV05/2019 | 45 - BetaCoV/Zhejiang/Hangzhou- 1/2020 | 625 |
| 64 - BetaCoV/Japan/AI/1-004/2020 | 79 - BetaCoV/Australia/QLD02/2020 | 619 |
| 19 - BetaCov/Wuhan/WH04/2020 | 45 - BetaCoV/Zhejiang/Hangzhou- 1/2020 | 618 |
| 8 - BetaCoV/Wuhan/WIV06/2019 | 50 - BetaCoV/Guangdong/20SF174/2020 | 616 |
| 2 - BetaCov/Wuhan/WHO01/2019 | 17 - BetaCoV/China/WHU01/2020 | 613 |
| 43 - BetaCoV/Shandong/IVDC-SD-00 1/2020 | 44 - BetaCoV/Jiangsu/IVDC-JS-00 1/2020 | 608 |
| 57 - BetaCoV/Guangdong/20SF201/2020 | 80 - BetaCoV/Taiwan/NTU01/2020 | 607 |
| 16 - BetaCov/Wuhan/WH03/2020 | 36 - BetaCoV/Zhejiang/WZ-02/2020 | 603 |
| 59 - BetaCoV/France/IDF0373/2020 | 68 - BetaCoV/Singapore/2/2020 | 602 |
| 34 - BetaCoV/Shenzhen/SZTH-004/2020 | 40 - BetaCoV/Beijing/IV DC-BJ-005/2020 | 598 |
| 36 - BetaCoV/Zhejiang/WZ-02/2020 | 80 - BetaCoV/Taiwan/NTU01/2020 | 597 |
| 16 - BetaCov/Wuhan/WH03/2020 | 33 - BetaCoV/Shenzhen/SZTH-003/2020 | 596 |
| 39 - BetaCoV/Chongqing/IVDC-CQ-00 1/2020 | 40 - BetaCoV/Beijing/IV DC-BJ-005/2020 | 595 |
| 16 - BetaCov/Wuhan/WH03/2020 | 50 - BetaCoV/Guangdong/20SF174/2020 | 595 |
| 8 - BetaCoV/Wuhan/WIV06/2019 | 21 - BetaCoV/Shenzhen/HKU-SZ-002/2020 | 594 |
| 44 - BetaCoV/Jiangsu/IVDC-JS-00 1/2020 | 46 - BetaCoV/USA/IL 1/2020 | 592 |
| 40 - BetaCoV/Beijing/IV DC-BJ-005/2020 | 41 - BetaCoV/USA/WA 1/2020 | 591 |
| 79 - BetaCoV/Australia/QLD02/2020 | 80 - BetaCoV/Taiwan/NTU01/2020 | 590 |
| 64 - BetaCoV/Japan/AI/1-004/2020 | 65 - BetaCoV/Korea/KCDC03/2020 | 588 |
| 6 - BetaCoV/Wuhan/WIV02/2019 | 19 - BetaCov/Wuhan/WH04/2020 | 587 |
| 8 - BetaCoV/Wuhan/WIV06/2019 | 36 - BetaCoV/Zhejiang/WZ-02/2020 | 587 |
| 9 - BetaCoV/Wuhan/WIV07/2019 | 65 - BetaCoV/Korea/KCDC03/2020 | 587 |
| 42 - BetaCoV/Hangzhou/HZCDC000 1/2020 | 79 - BetaCoV/Australia/QLD02/2020 | 587 |
| 6 - BetaCoV/Wuhan/WIV02/2019 | 42 - BetaCoV/Hangzhou/HZCDC000 1/2020 | 586 |
| 2 - BetaCov/Wuhan/WHO01/2019 | 32 - BetaCoV/Zhejiang/WZ-0 1/2020 | 585 |
| 19 - BetaCov/Wuhan/WH04/2020 | 79 - BetaCoV/Australia/QLD02/2020 | 585 |
| 8 - BetaCoV/Wuhan/WIV06/2019 | 33 - BetaCoV/Shenzhen/SZTH-003/2020 | 584 |
| 55 - BetaCoV/Taiwan/2/2020 | 58 - BetaCoV/France/IDF0372/2020 | 584 |
| 7 - BetaCoV/Wuhan/WIV05/2019 | 62 - BetaCoV/Australia/NS WO 1/2020 | 584 |
| 34 - BetaCoV/Shenzhen/SZTH-004/2020 | 50 - BetaCoV/Guangdong/20SF174/2020 | 583 |
| 44 - BetaCoV/Jiangsu/IVDC-JS-00 1/2020 | 59 - BetaCoV/France/IDF0373/2020 | 581 |
| 76 - BetaCoV/USA/CA5/2020 | 77 - BetaCoV/France/IDF05 15/2020 | 580 |
| 8 - BetaCoV/Wuhan/WIV06/2019 | 9 - BetaCoV/Wuhan/WIV07/2019 | 579 |
| 67 - BetaCoV/USA/W A 1-F6/2020 | 68 - BetaCoV/Singapore/2/2020 | 579 |
| 19 - BetaCov/Wuhan/WH04/2020 | 62 - BetaCoV/Australia/NS WO 1/2020 | 574 |
| 15 - BetaCoV/Wuhan/IPBCAMS-WH-05/2020 | 17 - BetaCoV/China/WHU01/2020 | 571 |
| 57 - BetaCoV/Guangdong/20SF201/2020 | 81 - BetaCoV/Singapore/3/2020 | 570 |
| 19 - BetaCov/Wuhan/WH04/2020 | 64 - BetaCoV/Japan/AI/1-004/2020 | 557 |
| 68 - BetaCoV/Singapore/2/2020 | 69 - BetaCoV/Germany/BavPat 1/2020 | 557 |
| 70 - BetaCoV/Australia/QLD001/2020 | 79 - BetaCoV/Australia/QLD02/2020 | 556 |
| 44 - BetaCoV/Jiangsu/IVDC-JS-00 1/2020 | 50 - BetaCoV/Guangdong/20SF174/2020 | 551 |
| 19 - BetaCov/Wuhan/WH04/2020 | 33 - BetaCoV/Shenzhen/SZTH-003/2020 | 550 |

**Table 4.** Edge weight between distinct nucleotide sequences in the Transcendental Information Cascade we constructed (contd.).

| Source sequence | Target sequence | Edge weight |
| --- | --- | --- |
| 23 - BetaCoV/Jiangxi/IV DC-JX-002/2020 | 24 - BetaCoV/Nonthaburi/74/2020 | 546 |
| 58 - BetaCoV/France/IDF0372/2020 | 59 - BetaCoV/France/IDF0373/2020 | 546 |
| 77 - BetaCoV/France/IDF05 15/2020 | 78 - BetaCoV/France/IDF0626/2020 | 545 |
| 10 - BetaCoV/Wuhan/HBCDC-HB-01/2019 | 11 - BetaCoV/Wuhan/IPBCAMS-WH-04/2019 | 544 |
| 24 - BetaCoV/Nonthaburi/74/2020 | 25 - BetaCoV/Shenzhen/SZTH-00 1/2020 | 544 |
| 26 - BetaCoV/Shenzhen/SZTH-002/2020 | 27 - BetaCoV/Guangdong/20SFO0 12/2020 | 544 |
| 30 - BetaCoV/Guangdong/20SF025/2020 | 31 - BetaCoV/Sichuan/ITVDC-SC-00 1/2020 | 544 |
| 37 - BetaCoV/Yunnan/IVDC- Y N-003/2020 | 38 - BetaCoV/Guangdong/20SF040/2020 | 544 |
| 38 - BetaCoV/Guangdong/20SF040/2020 | 39 - BetaCoV/Chongqing/IVDC-CQ-00 1/2020 | 544 |
| 59 - BetaCoV/France/IDF0373/2020 | 62 - BetaCoV/Australia/NS WO 1/2020 | 544 |
| 35 - BetaCoV/Guangdong/20SF028/2020 | 37 - BetaCoV/Yunnan/IVDC- Y N-003/2020 | 540 |
| 19 - BetaCoV/Wuhan/WH04/2020 | 21 - BetaCoV/Shenzhen/HKU-SZ-002/2020 | 539 |
| 77 - BetaCoV/France/IDF05 15/2020 | 81 - BetaCoV/Singapore/3/2020 | 539 |
| 6 - BetaCoV/Wuhan/WIV02/2019 | 10 - BetaCoV/Wuhan/HBCDC-HB-01/2019 | 538 |
| 7 - BetaCoV/Wuhan/WIV05/2019 | 64 - BetaCoV/Japan/A1/I-004/2020 | 536 |
| 2 - BetaCoV/Wuhan/WHO01/2019 | 65 - BetaCoV/Korea/KCDC03/2020 | 536 |
| 2 - BetaCoV/Wuhan/WHO01/2019 | 3 - BetaCoV/Wuhan/IV DC-HB-01/2019 | 535 |
| 54 - BetaCoV/Foshan/20SF21 1/2020 | 55 - BetaCoV/Taiwan/2/2020 | 535 |
| 1 - BetaCoV/Wuhan/IPBCAMS-WH-01/2019 | 2 - BetaCoV/Wuhan/WHO01/2019 | 534 |
| 18 - BetaCoV/China/WHU02/2020 | 20 - BetaCoV/Nonthaburi/6 1/2020 | 534 |
| 68 - BetaCoV/Singapore/2/2020 | 71 - BetaCoV/England/01/2020 | 534 |
| 7 - BetaCoV/Wuhan/WIV05/2019 | 42 - BetaCoV/Hangzhou/HZCDC000 1/2020 | 533 |
| 49 - BetaCoV/USA/AZ1/2020 | 50 - BetaCoV/Guangdong/20SF174/2020 | 533 |
| 15 - BetaCoV/Wuhan/IPBCAMS-WH-05/2020 | 16 - BetaCoV/Wuhan/WH03/2020 | 532 |
| 20 - BetaCoV/Nonthaburi/6 1/2020 | 22 - BetaCoV/Shenzhen/HKU-SZ-005/2020 | 532 |
| 21 - BetaCoV/Shenzhen/HKU-SZ-002/2020 | 22 - BetaCoV/Shenzhen/HKU-SZ-005/2020 | 532 |
| 56 - BetaCoV/USA/CA1/2020 | 57 - BetaCoV/Guangdong/20SF201/2020 | 532 |
| 30 - BetaCoV/Guangdong/20SF025/2020 | 32 - BetaCoV/Zhejiang/WZ-0 1/2020 | 531 |
| 2 - BetaCoV/Wuhan/WHO01/2019 | 6 - BetaCoV/Wuhan/WIV02/2019 | 530 |
| 56 - BetaCoV/USA/CA1/2020 | 58 - BetaCoV/France/IDF0372/2020 | 530 |
| 58 - BetaCoV/France/IDF0372/2020 | 60 - BetaCoV/Singapore/ 1/2020 | 529 |
| 69 - BetaCoV/Germany/BavPat 1/2020 | 77 - BetaCoV/France/IDF05 15/2020 | 528 |
| 55 - BetaCoV/Taiwan/2/2020 | 57 - BetaCoV/Guangdong/20SF201/2020 | 526 |
| 57 - BetaCoV/Guangdong/20SF201/2020 | 68 - BetaCoV/Singapore/2/2020 | 526 |
| 80 - BetaCoV/Taiwan/NTU01/2020 | 82 - BetaCoV/Belgium/GHB-0302 1/2020 | 526 |
| 18 - BetaCoV/China/WHU02/2020 | 22 - BetaCoV/Shenzhen/HKU-SZ-005/2020 | 525 |
| 70 - BetaCoV/Australia/QLD001/2020 | 78 - BetaCoV/France/IDF0626/2020 | 525 |
| 76 - BetaCoV/USA/CA5/2020 | 78 - BetaCoV/France/IDF0626/2020 | 525 |
| 18 - BetaCoV/China/WHU02/2020 | 19 - BetaCoV/Wuhan/WH04/2020 | 524 |
| 70 - BetaCoV/Australia/QLD001/2020 | 80 - BetaCoV/Taiwan/NTU01/2020 | 524 |
| 38 - BetaCoV/Guangdong/20SF040/2020 | 40 - BetaCoV/Beijing/IV DC-BJ-005/2020 | 523 |
| 45 - BetaCoV/Zhejiang/Hangzhou- 1/2020 | 50 - BetaCoV/Guangdong/20SF174/2020 | 522 |
| 54 - BetaCoV/Foshan/20SF21 1/2020 | 56 - BetaCoV/USA/CA1/2020 | 522 |
| 44 - BetaCoV/Jiangsu/IVDC-JS-00 1/2020 | 65 - BetaCoV/Korea/KCDC03/2020 | 521 |
| 34 - BetaCoV/Shenzhen/SZTH-004/2020 | 80 - BetaCoV/Taiwan/NTU01/2020 | 519 |
| 32 - BetaCoV/Zhejiang/WZ-0 1/2020 | 35 - BetaCoV/Guangdong/20SF028/2020 | 518 |
| 62 - BetaCoV/Australia/NS WO 1/2020 | 68 - BetaCoV/Singapore/2/2020 | 517 |
| 2 - BetaCoV/Wuhan/WHO01/2019 | 8 - BetaCoV/Wuhan/WIV06/2019 | 516 |
| 31 - BetaCoV/Sichuan/ITVDC-SC-00 1/2020 | 35 - BetaCoV/Guangdong/20SF028/2020 | 516 |
| 6 - BetaCoV/Wuhan/WIV02/2019 | 45 - BetaCoV/Zhejiang/Hangzhou- 1/2020 | 516 |
| 38 - BetaCoV/Guangdong/20SF040/2020 | 45 - BetaCoV/Zhejiang/Hangzhou- 1/2020 | 516 |
| 58 - BetaCoV/France/IDF0372/2020 | 62 - BetaCoV/Australia/NS WO 1/2020 | 515 |
| 69 - BetaCoV/Germany/BavPat 1/2020 | 78 - BetaCoV/France/IDF0626/2020 | 514 |
| 59 - BetaCoV/France/IDF0373/2020 | 64 - BetaCoV/Japan/A1/I-004/2020 | 512 |
| 70 - BetaCoV/Australia/QLD001/2020 | 71 - BetaCoV/England/01/2020 | 512 |
| 10 - BetaCoV/Wuhan/HBCDC-HB-01/2019 | 17 - BetaCoV/China/WHU01/2020 | 511 |
| 65 - BetaCoV/Korea/KCDC03/2020 | 66 - BetaCoV/USA/WA1-A12/2020 | 511 |
| 57 - BetaCoV/Guangdong/20SF201/2020 | 60 - BetaCoV/Singapore/ 1/2020 | 510 |
| 42 - BetaCoV/Hangzhou/HZCDC000 1/2020 | 44 - BetaCoV/Jiangsu/IVDC-JS-00 1/2020 | 509 |
| 21 - BetaCoV/Shenzhen/HKU-SZ-002/2020 | 80 - BetaCoV/Taiwan/NTU01/2020 | 509 |
| 40 - BetaCoV/Beijing/IV DC-BJ-005/2020 | 42 - BetaCoV/Hangzhou/HZCDC000 1/2020 | 508 |
| 55 - BetaCoV/Taiwan/2/2020 | 79 - BetaCoV/Australia/QLD02/2020 | 507 |
| 31 - BetaCoV/Sichuan/ITVDC-SC-00 1/2020 | 33 - BetaCoV/Shenzhen/SZTH-003/2020 | 505 |
| 16 - BetaCoV/Wuhan/WH03/2020 | 80 - BetaCoV/Taiwan/NTU01/2020 | 505 |
| 78 - BetaCoV/France/IDF0626/2020 | 82 - BetaCoV/Belgium/GHB-0302 1/2020 | 505 |
| 34 - BetaCoV/Shenzhen/SZTH-004/2020 | 37 - BetaCoV/Yunnan/IVDC- Y N-003/2020 | 504 |
| 18 - BetaCoV/China/WHU02/2020 | 21 - BetaCoV/Shenzhen/HKU-SZ-002/2020 | 503 |
| 7 - BetaCoV/Wuhan/WIV05/2019 | 79 - BetaCoV/Australia/QLD02/2020 | 502 |
| 16 - BetaCoV/Wuhan/WH03/2020 | 32 - BetaCoV/Zhejiang/WZ-0 1/2020 | 501 |
| 5 - BetaCoV/Wuhan/WIV04/2019 | 8 - BetaCoV/Wuhan/WIV06/2019 | 497 |
| 16 - BetaCoV/Wuhan/WH03/2020 | 17 - BetaCoV/China/WHU01/2020 | 497 |
| 15 - BetaCoV/Wuhan/IPBCAMS-WH-05/2020 | 20 - BetaCoV/Nonthaburi/6 1/2020 | 497 |
| 31 - BetaCoV/Sichuan/ITVDC-SC-00 1/2020 | 32 - BetaCoV/Zhejiang/WZ-0 1/2020 | 497 |
| 59 - BetaCoV/France/IDF0373/2020 | 60 - BetaCoV/Singapore/ 1/2020 | 496 |
| 76 - BetaCoV/USA/CA5/2020 | 80 - BetaCoV/Taiwan/NTU01/2020 | 494 |
| 19 - BetaCoV/Wuhan/WH04/2020 | 20 - BetaCoV/Nonthaburi/6 1/2020 | 493 |
| 32 - BetaCoV/Zhejiang/WZ-0 1/2020 | 42 - BetaCoV/Hangzhou/HZCDC000 1/2020 | 493 |
| 58 - BetaCoV/France/IDF0372/2020 | 65 - BetaCoV/Korea/KCDC03/2020 | 493 |
| 63 - BetaCoV/Australia/VIC01/2020 | 65 - BetaCoV/Korea/KCDC03/2020 | 492 |
| 6 - BetaCoV/Wuhan/WIV02/2019 | 9 - BetaCoV/Wuhan/WIV07/2019 | 491 |
| 34 - BetaCoV/Shenzhen/SZTH-004/2020 | 42 - BetaCoV/Hangzhou/HZCDC000 1/2020 | 489 |
| 8 - BetaCoV/Wuhan/WIV06/2019 | 11 - BetaCoV/Wuhan/IPBCAMS-WH-04/2019 | 488 |
| 32 - BetaCoV/Zhejiang/WZ-0 1/2020 | 33 - BetaCoV/Shenzhen/SZTH-003/2020 | 488 |
| 36 - BetaCoV/Zhejiang/WZ-02/2020 | 37 - BetaCoV/Yunnan/IVDC- Y N-003/2020 | 488 |
| 36 - BetaCoV/Zhejiang/WZ-02/2020 | 40 - BetaCoV/Beijing/IV DC-BJ-005/2020 | 488 |
| 69 - BetaCoV/Germany/BavPat 1/2020 | 70 - BetaCoV/Australia/QLD001/2020 | 487 |
| 15 - BetaCoV/Wuhan/IPBCAMS-WH-05/2020 | 21 - BetaCoV/Shenzhen/HKU-SZ-002/2020 | 485 |
| 42 - BetaCoV/Hangzhou/HZCDC000 1/2020 | 59 - BetaCoV/France/IDF0373/2020 | 485 |
| 2 - BetaCoV/Wuhan/WHO01/2019 | 10 - BetaCoV/Wuhan/HBCDC-HB-01/2019 | 484 |
| 7 - BetaCoV/Wuhan/WIV05/2019 | 16 - BetaCoV/Wuhan/WH03/2020 | 483 |
| 34 - BetaCoV/Shenzhen/SZTH-004/2020 | 81 - BetaCoV/Singapore/3/2020 | 483 |
| 5 - BetaCoV/Wuhan/WIV04/2019 | 10 - BetaCoV/Wuhan/HBCDC-HB-01/2019 | 482 |
| 9 - BetaCoV/Wuhan/WIV07/2019 | 11 - BetaCoV/Wuhan/IPBCAMS-WH-04/2019 | 482 |
| 7 - BetaCoV/Wuhan/WIV05/2019 | 17 - BetaCoV/China/WHU01/2020 | 482 |
| 59 - BetaCoV/France/IDF0373/2020 | 78 - BetaCoV/France/IDF0626/2020 | 482 |
| 16 - BetaCoV/Wuhan/WH03/2020 | 19 - BetaCoV/Wuhan/WH04/2020 | 481 |
| 18 - BetaCoV/China/WHU02/2020 | 33 - BetaCoV/Shenzhen/SZTH-003/2020 | 481 |
| 19 - BetaCoV/Wuhan/WH04/2020 | 36 - BetaCoV/Zhejiang/WZ-02/2020 | 481 |
| 67 - BetaCoV/USA/W A 1-F6/2020 | 70 - BetaCoV/Australia/QLD001/2020 | 481 |
| 6 - BetaCoV/Wuhan/WIV02/2019 | 8 - BetaCoV/Wuhan/WIV06/2019 | 480 |

**Table 5.**
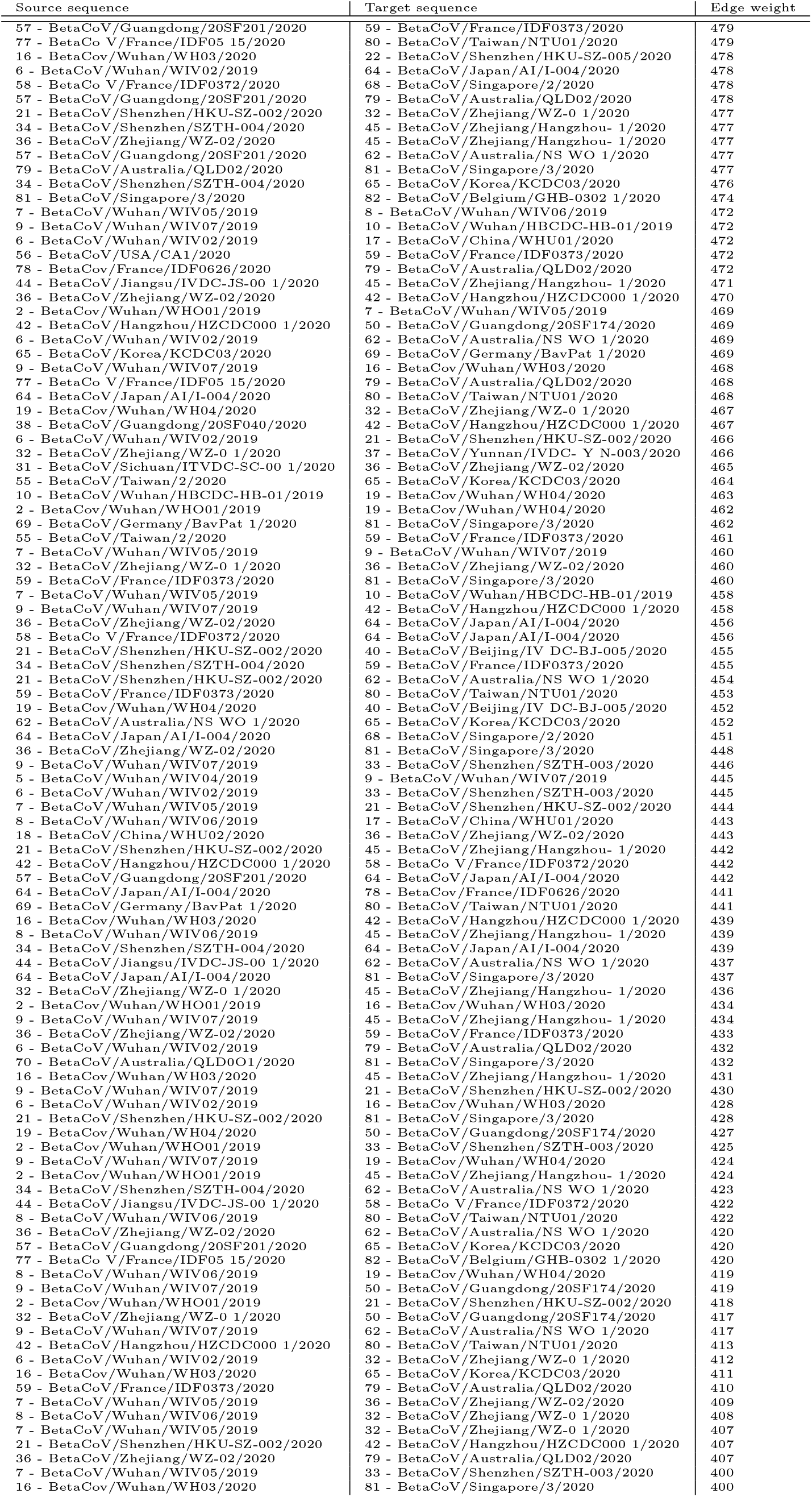
Edge weight between distinct nucleotide sequences in the Transcendental Information Cascade we constructed.

**Table 6.**
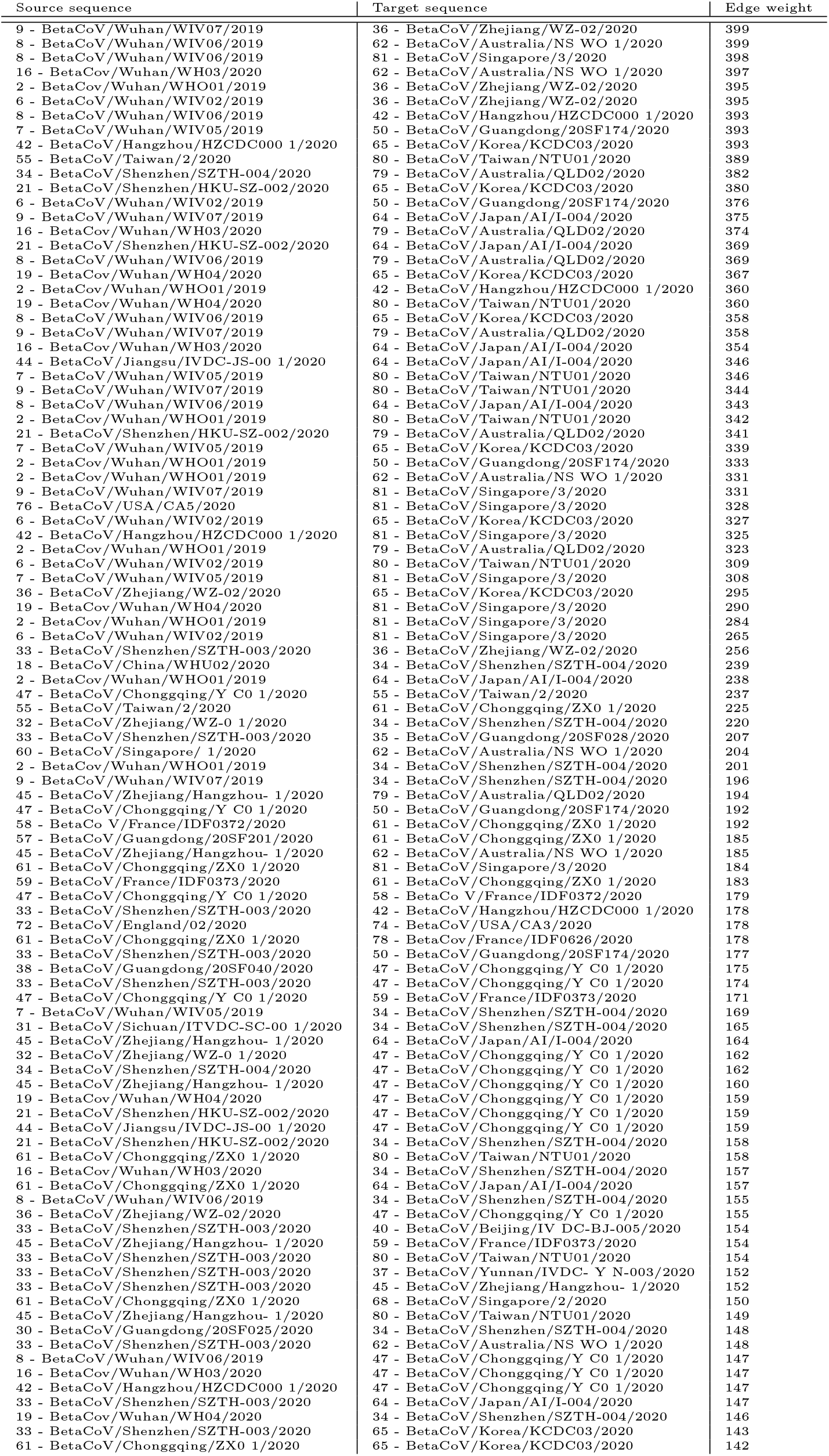
Edge weight between distinct nucleotide sequences in the Transcendental Information Cascade we constructed.

| Source sequence | Target sequence | Edge weight |
| --- | --- | --- |
| 9 - BetaCoV/Wuhan/WIV07/2019 | 36 - BetaCoV/Zhejiang/WZ-02/2020 | 399 |
| 8 - BetaCoV/Wuhan/WIV06/2019 | 62 - BetaCoV/Australia/NS WO 1/2020 | 399 |
| 8 - BetaCoV/Wuhan/WIV06/2019 | 81 - BetaCoV/Singapore/3/2020 | 398 |
| 16 - BetaCov/Wuhan/WH03/2020 | 62 - BetaCoV/Australia/NS WO 1/2020 | 397 |
| 2 - BetaCov/Wuhan/WHO01/2019 | 36 - BetaCoV/Zhejiang/WZ-02/2020 | 395 |
| 6 - BetaCoV/Wuhan/WIV02/2019 | 36 - BetaCoV/Zhejiang/WZ-02/2020 | 395 |
| 8 - BetaCoV/Wuhan/WIV06/2019 | 42 - BetaCoV/Hangzhou/HZCDC000 1/2020 | 393 |
| 7 - BetaCoV/Wuhan/WIV05/2019 | 50 - BetaCoV/Guangdong/20SF174/2020 | 393 |
| 42 - BetaCoV/Hangzhou/HZCDC000 1/2020 | 65 - BetaCoV/Korea/KCDC03/2020 | 393 |
| 55 - BetaCoV/Taiwan/2/2020 | 80 - BetaCoV/Taiwan/NTU01/2020 | 389 |
| 34 - BetaCoV/Shenzhen/SZTH-004/2020 | 79 - BetaCoV/Australia/QLD02/2020 | 382 |
| 21 - BetaCoV/Shenzhen/HKU-SZ-002/2020 | 65 - BetaCoV/Korea/KCDC03/2020 | 380 |
| 6 - BetaCoV/Wuhan/WIV02/2019 | 50 - BetaCoV/Guangdong/20SF174/2020 | 376 |
| 9 - BetaCoV/Wuhan/WIV07/2019 | 64 - BetaCoV/Japan/AI/I-004/2020 | 375 |
| 16 - BetaCov/Wuhan/WH03/2020 | 79 - BetaCoV/Australia/QLD02/2020 | 374 |
| 21 - BetaCoV/Shenzhen/HKU-SZ-002/2020 | 64 - BetaCoV/Japan/AI/I-004/2020 | 369 |
| 8 - BetaCoV/Wuhan/WIV06/2019 | 79 - BetaCoV/Australia/QLD02/2020 | 369 |
| 19 - BetaCov/Wuhan/WH04/2020 | 65 - BetaCoV/Korea/KCDC03/2020 | 367 |
| 2 - BetaCov/Wuhan/WHO01/2019 | 42 - BetaCoV/Hangzhou/HZCDC000 1/2020 | 360 |
| 19 - BetaCov/Wuhan/WH04/2020 | 80 - BetaCoV/Taiwan/NTU01/2020 | 360 |
| 8 - BetaCoV/Wuhan/WIV06/2019 | 65 - BetaCoV/Korea/KCDC03/2020 | 358 |
| 9 - BetaCoV/Wuhan/WIV07/2019 | 79 - BetaCoV/Australia/QLD02/2020 | 358 |
| 16 - BetaCov/Wuhan/WH03/2020 | 64 - BetaCoV/Japan/AI/I-004/2020 | 354 |
| 44 - BetaCoV/Jiangsu/IVDC-JS-00 1/2020 | 64 - BetaCoV/Japan/AI/I-004/2020 | 346 |
| 7 - BetaCoV/Wuhan/WIV05/2019 | 80 - BetaCoV/Taiwan/NTU01/2020 | 346 |
| 9 - BetaCoV/Wuhan/WIV07/2019 | 80 - BetaCoV/Taiwan/NTU01/2020 | 344 |
| 8 - BetaCoV/Wuhan/WIV06/2019 | 64 - BetaCoV/Japan/AI/I-004/2020 | 343 |
| 2 - BetaCov/Wuhan/WHO01/2019 | 80 - BetaCoV/Taiwan/NTU01/2020 | 342 |
| 21 - BetaCoV/Shenzhen/HKU-SZ-002/2020 | 79 - BetaCoV/Australia/QLD02/2020 | 341 |
| 7 - BetaCoV/Wuhan/WIV05/2019 | 65 - BetaCoV/Korea/KCDC03/2020 | 339 |
| 2 - BetaCov/Wuhan/WHO01/2019 | 50 - BetaCoV/Guangdong/20SF174/2020 | 333 |
| 2 - BetaCov/Wuhan/WHO01/2019 | 62 - BetaCoV/Australia/NS WO 1/2020 | 331 |
| 9 - BetaCoV/Wuhan/WIV07/2019 | 81 - BetaCoV/Singapore/3/2020 | 331 |
| 76 - BetaCoV/USA/CA5/2020 | 81 - BetaCoV/Singapore/3/2020 | 328 |
| 6 - BetaCoV/Wuhan/WIV02/2019 | 65 - BetaCoV/Korea/KCDC03/2020 | 327 |
| 42 - BetaCoV/Hangzhou/HZCDC000 1/2020 | 81 - BetaCoV/Singapore/3/2020 | 325 |
| 2 - BetaCov/Wuhan/WHO01/2019 | 79 - BetaCoV/Australia/QLD02/2020 | 323 |
| 6 - BetaCoV/Wuhan/WIV02/2019 | 80 - BetaCoV/Taiwan/NTU01/2020 | 309 |
| 7 - BetaCoV/Wuhan/WIV05/2019 | 81 - BetaCoV/Singapore/3/2020 | 308 |
| 36 - BetaCoV/Zhejiang/WZ-02/2020 | 65 - BetaCoV/Korea/KCDC03/2020 | 295 |
| 19 - BetaCov/Wuhan/WH04/2020 | 81 - BetaCoV/Singapore/3/2020 | 290 |
| 2 - BetaCov/Wuhan/WHO01/2019 | 81 - BetaCoV/Singapore/3/2020 | 284 |
| 6 - BetaCoV/Wuhan/WIV02/2019 | 81 - BetaCoV/Singapore/3/2020 | 265 |
| 33 - BetaCoV/Shenzhen/SZTH-003/2020 | 36 - BetaCoV/Zhejiang/WZ-02/2020 | 256 |
| 18 - BetaCoV/China/WHU02/2020 | 34 - BetaCoV/Shenzhen/SZTH-004/2020 | 239 |
| 2 - BetaCov/Wuhan/WHO01/2019 | 64 - BetaCoV/Japan/AI/I-004/2020 | 238 |
| 47 - BetaCoV/Chongqing/Y C0 1/2020 | 55 - BetaCoV/Taiwan/2/2020 | 237 |
| 55 - BetaCoV/Taiwan/2/2020 | 61 - BetaCoV/Chongqing/ZX0 1/2020 | 225 |
| 32 - BetaCoV/Zhejiang/WZ-0 1/2020 | 34 - BetaCoV/Shenzhen/SZTH-004/2020 | 220 |
| 33 - BetaCoV/Shenzhen/SZTH-003/2020 | 35 - BetaCoV/Guangdong/20SF028/2020 | 207 |
| 60 - BetaCoV/Singapore/ 1/2020 | 62 - BetaCoV/Australia/NS WO 1/2020 | 204 |
| 2 - BetaCoV/Wuhan/WHO01/2019 | 34 - BetaCoV/Shenzhen/SZTH-004/2020 | 201 |
| 9 - BetaCoV/Wuhan/WIV07/2019 | 34 - BetaCoV/Shenzhen/SZTH-004/2020 | 196 |
| 45 - BetaCoV/Zhejiang/Hangzhou- 1/2020 | 79 - BetaCoV/Australia/QLD02/2020 | 194 |
| 47 - BetaCoV/Chongqing/Y C0 1/2020 | 50 - BetaCoV/Guangdong/20SF174/2020 | 192 |
| 58 - BetaCoV/France/IDF0372/2020 | 61 - BetaCoV/Chongqing/ZX0 1/2020 | 192 |
| 57 - BetaCoV/Guangdong/20SF201/2020 | 61 - BetaCoV/Chongqing/ZX0 1/2020 | 185 |
| 45 - BetaCoV/Zhejiang/Hangzhou- 1/2020 | 62 - BetaCoV/Australia/NS WO 1/2020 | 185 |
| 61 - BetaCoV/Chongqing/ZX0 1/2020 | 81 - BetaCoV/Singapore/3/2020 | 184 |
| 59 - BetaCoV/France/IDF0373/2020 | 61 - BetaCoV/Chongqing/ZX0 1/2020 | 183 |
| 47 - BetaCoV/Chongqing/Y C0 1/2020 | 58 - BetaCoV/France/IDF0372/2020 | 179 |
| 33 - BetaCoV/Shenzhen/SZTH-003/2020 | 42 - BetaCoV/Hangzhou/HZCDC000 1/2020 | 178 |
| 72 - BetaCoV/England/02/2020 | 74 - BetaCoV/USA/CA3/2020 | 178 |
| 61 - BetaCoV/Chongqing/ZX0 1/2020 | 78 - BetaCoV/France/IDF0626/2020 | 178 |
| 33 - BetaCoV/Shenzhen/SZTH-003/2020 | 50 - BetaCoV/Guangdong/20SF174/2020 | 177 |
| 38 - BetaCoV/Guangdong/20SF040/2020 | 47 - BetaCoV/Chongqing/Y C0 1/2020 | 175 |
| 33 - BetaCoV/Shenzhen/SZTH-003/2020 | 47 - BetaCoV/Chongqing/Y C0 1/2020 | 174 |
| 47 - BetaCoV/Chongqing/Y C0 1/2020 | 59 - BetaCoV/France/IDF0373/2020 | 171 |
| 7 - BetaCoV/Wuhan/WIV05/2019 | 34 - BetaCoV/Shenzhen/SZTH-004/2020 | 169 |
| 31 - BetaCoV/Sichuan/ITVDC-SC-00 1/2020 | 34 - BetaCoV/Shenzhen/SZTH-004/2020 | 165 |
| 45 - BetaCoV/Zhejiang/Hangzhou- 1/2020 | 64 - BetaCoV/Japan/AI/I-004/2020 | 164 |
| 32 - BetaCoV/Zhejiang/WZ-0 1/2020 | 47 - BetaCoV/Chongqing/Y C0 1/2020 | 162 |
| 34 - BetaCoV/Shenzhen/SZTH-004/2020 | 47 - BetaCoV/Chongqing/Y C0 1/2020 | 162 |
| 45 - BetaCoV/Zhejiang/Hangzhou- 1/2020 | 47 - BetaCoV/Chongqing/Y C0 1/2020 | 160 |
| 19 - BetaCov/Wuhan/WH04/2020 | 47 - BetaCoV/Chongqing/Y C0 1/2020 | 159 |
| 21 - BetaCoV/Shenzhen/HKU-SZ-002/2020 | 47 - BetaCoV/Chongqing/Y C0 1/2020 | 159 |
| 44 - BetaCoV/Jiangsu/IVDC-JS-00 1/2020 | 47 - BetaCoV/Chongqing/Y C0 1/2020 | 159 |
| 21 - BetaCoV/Shenzhen/HKU-SZ-002/2020 | 34 - BetaCoV/Shenzhen/SZTH-004/2020 | 158 |
| 61 - BetaCoV/Chongqing/ZX0 1/2020 | 80 - BetaCoV/Taiwan/NTU01/2020 | 158 |
| 16 - BetaCov/Wuhan/WH03/2020 | 34 - BetaCoV/Shenzhen/SZTH-004/2020 | 157 |
| 61 - BetaCoV/Chongqing/ZX0 1/2020 | 64 - BetaCoV/Japan/AI/I-004/2020 | 157 |
| 8 - BetaCoV/Wuhan/WIV06/2019 | 34 - BetaCoV/Shenzhen/SZTH-004/2020 | 155 |
| 36 - BetaCoV/Zhejiang/WZ-02/2020 | 47 - BetaCoV/Chongqing/Y C0 1/2020 | 155 |
| 33 - BetaCoV/Shenzhen/SZTH-003/2020 | 40 - BetaCoV/Beijing/IV DC-BJ-005/2020 | 154 |
| 45 - BetaCoV/Zhejiang/Hangzhou- 1/2020 | 59 - BetaCoV/France/IDF0373/2020 | 154 |
| 33 - BetaCoV/Shenzhen/SZTH-003/2020 | 80 - BetaCoV/Taiwan/NTU01/2020 | 154 |
| 33 - BetaCoV/Shenzhen/SZTH-003/2020 | 37 - BetaCoV/Yunnan/IVDC- Y N-003/2020 | 152 |
| 33 - BetaCoV/Shenzhen/SZTH-003/2020 | 45 - BetaCoV/Zhejiang/Hangzhou- 1/2020 | 152 |
| 61 - BetaCoV/Chongqing/ZX0 1/2020 | 68 - BetaCoV/Singapore/2/2020 | 150 |
| 45 - BetaCoV/Zhejiang/Hangzhou- 1/2020 | 80 - BetaCoV/Taiwan/NTU01/2020 | 149 |
| 30 - BetaCoV/Guangdong/20SF025/2020 | 34 - BetaCoV/Shenzhen/SZTH-004/2020 | 148 |
| 33 - BetaCoV/Shenzhen/SZTH-003/2020 | 62 - BetaCoV/Australia/NS WO 1/2020 | 148 |
| 8 - BetaCoV/Wuhan/WIV06/2019 | 47 - BetaCoV/Chongqing/Y C0 1/2020 | 147 |
| 16 - BetaCov/Wuhan/WH03/2020 | 47 - BetaCoV/Chongqing/Y C0 1/2020 | 147 |
| 42 - BetaCoV/Hangzhou/HZCDC000 1/2020 | 47 - BetaCoV/Chongqing/Y C0 1/2020 | 147 |
| 33 - BetaCoV/Shenzhen/SZTH-003/2020 | 64 - BetaCoV/Japan/AI/I-004/2020 | 147 |
| 19 - BetaCov/Wuhan/WH04/2020 | 34 - BetaCoV/Shenzhen/SZTH-004/2020 | 146 |
| 33 - BetaCoV/Shenzhen/SZTH-003/2020 | 65 - BetaCoV/Korea/KCDC03/2020 | 143 |
| 61 - BetaCoV/Chongqing/ZX0 1/2020 | 65 - BetaCoV/Korea/KCDC03/2020 | 142 |

**Table 7.**
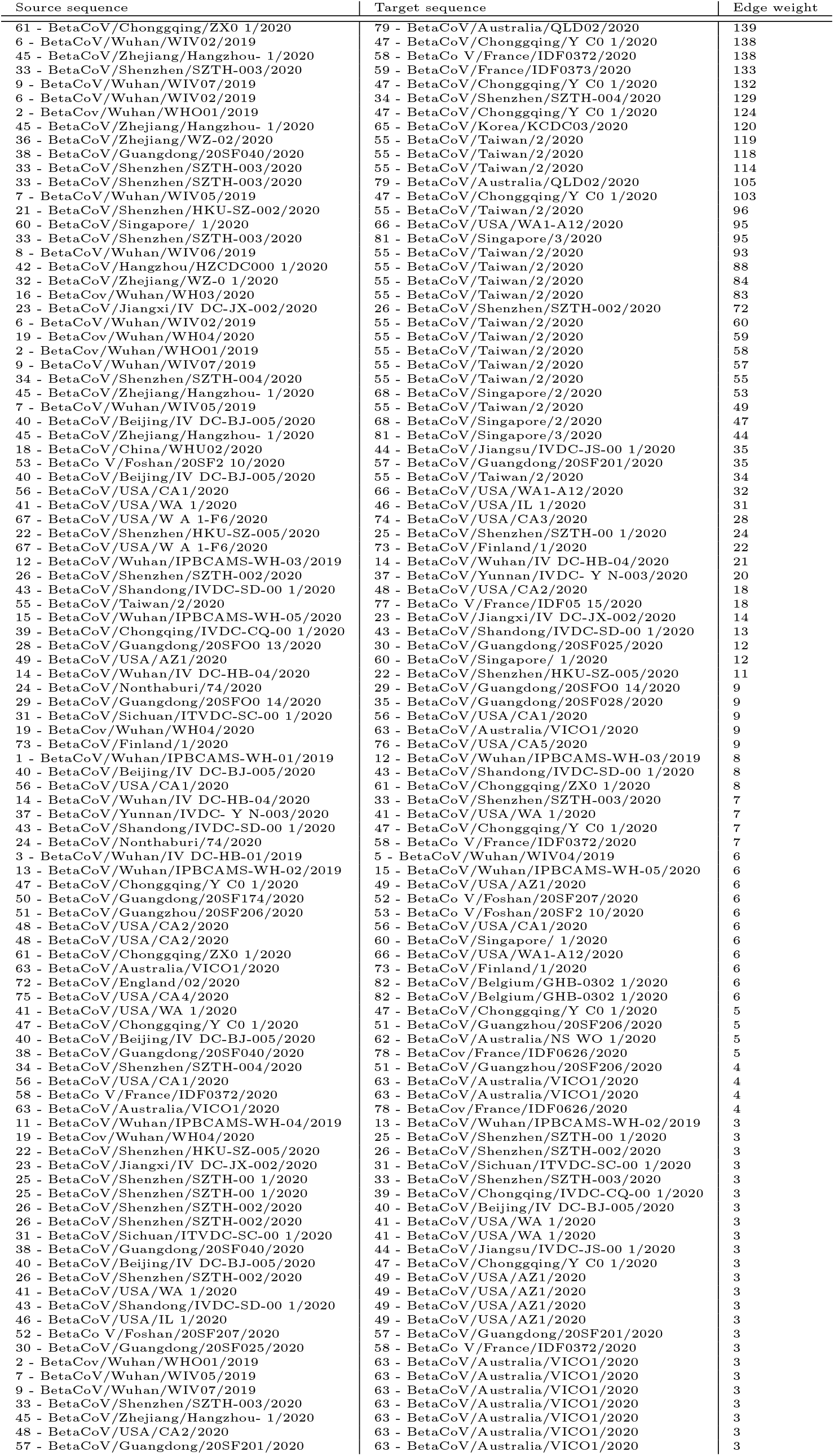
Edge weight between distinct nucleotide sequences in the Transcendental Information Cascade we constructed.

**Table 8.**
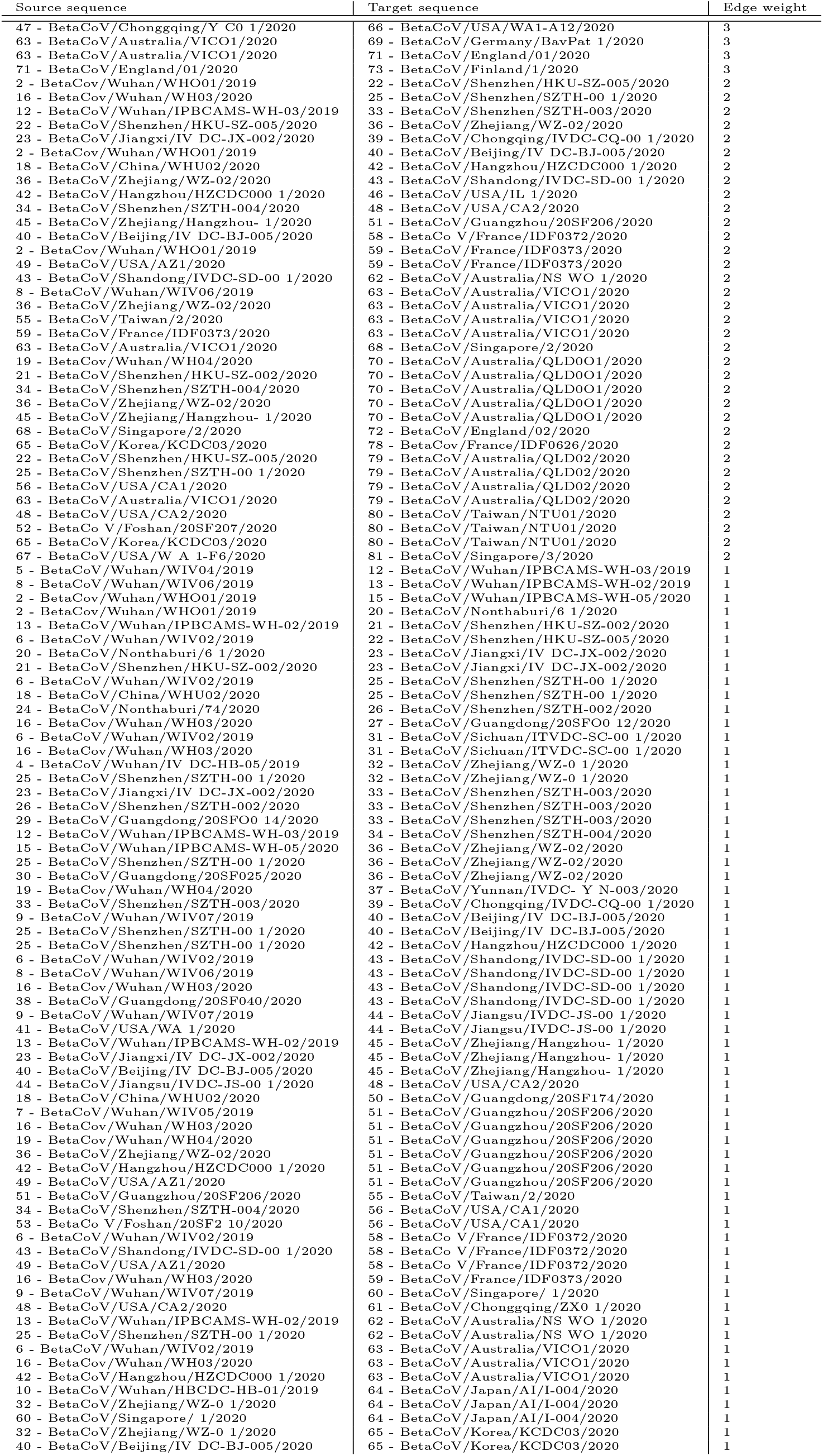
Edge weight between distinct nucleotide sequences in the Transcendental Information Cascade we constructed.

**Table 9.** Edge weight between distinct nucleotide sequences in the Transcendental Information Cascade we constructed.

| Source sequence | Target sequence | Edge weight |
| --- | --- | --- |
| 60 - BetaCoV/Singapore/ 1/2020 | 65 - BetaCoV/Korea/KCDC03/2020 | 1 |
| 58 - BetaCoV/France/IDF0372/2020 | 66 - BetaCoV/USA/WA1-A12/2020 | 1 |
| 62 - BetaCoV/Australia/NS WO 1/2020 | 66 - BetaCoV/USA/WA1-A12/2020 | 1 |
| 18 - BetaCoV/China/WHU02/2020 | 68 - BetaCoV/Singapore/2/2020 | 1 |
| 36 - BetaCoV/Zhejiang/WZ-02/2020 | 68 - BetaCoV/Singapore/2/2020 | 1 |
| 42 - BetaCoV/Hangzhou/HZCDC000 1/2020 | 68 - BetaCoV/Singapore/2/2020 | 1 |
| 2 - BetaCoV/Wuhan/WHO01/2019 | 69 - BetaCoV/Germany/BavPat 1/2020 | 1 |
| 16 - BetaCoV/Wuhan/WH03/2020 | 69 - BetaCoV/Germany/BavPat 1/2020 | 1 |
| 19 - BetaCoV/Wuhan/WH04/2020 | 69 - BetaCoV/Germany/BavPat 1/2020 | 1 |
| 57 - BetaCoV/Guangdong/20SF201/2020 | 69 - BetaCoV/Germany/BavPat 1/2020 | 1 |
| 2 - BetaCoV/Wuhan/WHO01/2019 | 70 - BetaCoV/Australia/QLD001/2020 | 1 |
| 7 - BetaCoV/Wuhan/WIV05/2019 | 70 - BetaCoV/Australia/QLD001/2020 | 1 |
| 8 - BetaCoV/Wuhan/WIV06/2019 | 70 - BetaCoV/Australia/QLD001/2020 | 1 |
| 33 - BetaCoV/Shenzhen/SZTH-003/2020 | 70 - BetaCoV/Australia/QLD001/2020 | 1 |
| 58 - BetaCoV/France/IDF0372/2020 | 70 - BetaCoV/Australia/QLD001/2020 | 1 |
| 61 - BetaCoV/Chongqing/ZX0 1/2020 | 70 - BetaCoV/Australia/QLD001/2020 | 1 |
| 64 - BetaCoV/Japan/AI/I-004/2020 | 70 - BetaCoV/Australia/QLD001/2020 | 1 |
| 34 - BetaCoV/Shenzhen/SZTH-004/2020 | 71 - BetaCoV/England/01/2020 | 1 |
| 45 - BetaCoV/Zhejiang/Hangzhou- 1/2020 | 71 - BetaCoV/England/01/2020 | 1 |
| 55 - BetaCoV/Taiwan/2/2020 | 71 - BetaCoV/England/01/2020 | 1 |
| 19 - BetaCoV/Wuhan/WH04/2020 | 72 - BetaCoV/England/02/2020 | 1 |
| 21 - BetaCoV/Shenzhen/HKU-SZ-002/2020 | 72 - BetaCoV/England/02/2020 | 1 |
| 67 - BetaCoV/USA/W A 1-F6/2020 | 72 - BetaCoV/England/02/2020 | 1 |
| 2 - BetaCoV/Wuhan/WHO01/2019 | 74 - BetaCoV/USA/CA3/2020 | 1 |
| 57 - BetaCoV/Guangdong/20SF201/2020 | 76 - BetaCoV/USA/CA5/2020 | 1 |
| 19 - BetaCoV/Wuhan/WH04/2020 | 77 - BetaCoV/France/IDF05 15/2020 | 1 |
| 58 - BetaCoV/France/IDF0372/2020 | 77 - BetaCoV/France/IDF05 15/2020 | 1 |
| 59 - BetaCoV/France/IDF0373/2020 | 77 - BetaCoV/France/IDF05 15/2020 | 1 |
| 61 - BetaCoV/Chongqing/ZX0 1/2020 | 77 - BetaCoV/France/IDF05 15/2020 | 1 |
| 55 - BetaCoV/Taiwan/2/2020 | 78 - BetaCoV/France/IDF0626/2020 | 1 |
| 14 - BetaCoV/Wuhan/IV DC-HB-04/2020 | 79 - BetaCoV/Australia/QLD02/2020 | 1 |
| 31 - BetaCoV/Sichuan/ITVDC-SC-00 1/2020 | 79 - BetaCoV/Australia/QLD02/2020 | 1 |
| 32 - BetaCoV/Zhejiang/WZ-0 1/2020 | 79 - BetaCoV/Australia/QLD02/2020 | 1 |
| 13 - BetaCoV/Wuhan/IPBCAMS-WH-02/2019 | 80 - BetaCoV/Taiwan/NTU01/2020 | 1 |
| 60 - BetaCoV/Singapore/ 1/2020 | 80 - BetaCoV/Taiwan/NTU01/2020 | 1 |

**Table 10.**
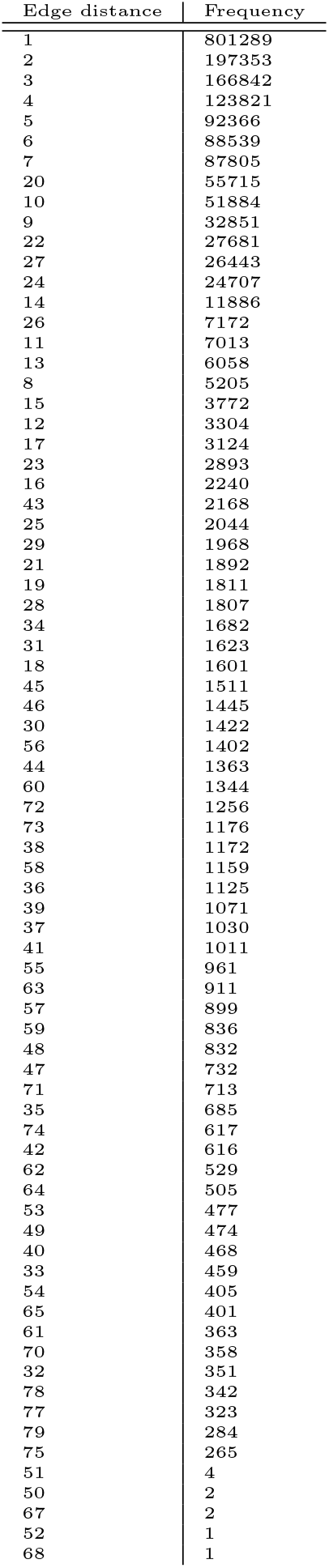
Frequency of edge distances.

| Edge distance | Frequency |
| --- | --- |
| 1 | 801289 |
| 2 | 197353 |
| 3 | 166842 |
| 4 | 123821 |
| 5 | 92366 |
| 6 | 88539 |
| 7 | 87805 |
| 20 | 55715 |
| 10 | 51884 |
| 9 | 32851 |
| 22 | 27681 |
| 27 | 26443 |
| 24 | 24707 |
| 14 | 11886 |
| 26 | 7172 |
| 11 | 7013 |
| 13 | 6058 |
| 8 | 5205 |
| 15 | 3772 |
| 12 | 3304 |
| 17 | 3124 |
| 23 | 2893 |
| 16 | 2240 |
| 43 | 2168 |
| 25 | 2044 |
| 29 | 1968 |
| 21 | 1892 |
| 19 | 1811 |
| 28 | 1807 |
| 34 | 1682 |
| 31 | 1623 |
| 18 | 1601 |
| 45 | 1511 |
| 46 | 1445 |
| 30 | 1422 |
| 56 | 1402 |
| 44 | 1363 |
| 60 | 1344 |
| 72 | 1256 |
| 73 | 1176 |
| 38 | 1172 |
| 58 | 1159 |
| 36 | 1125 |
| 39 | 1071 |
| 37 | 1030 |
| 41 | 1011 |
| 55 | 961 |
| 63 | 911 |
| 57 | 899 |
| 59 | 836 |
| 48 | 832 |
| 47 | 732 |
| 71 | 713 |
| 35 | 685 |
| 74 | 617 |
| 42 | 616 |
| 62 | 529 |
| 64 | 505 |
| 53 | 477 |
| 49 | 474 |
| 40 | 468 |
| 33 | 459 |
| 54 | 405 |
| 65 | 401 |
| 61 | 363 |
| 70 | 358 |
| 32 | 351 |
| 78 | 342 |
| 77 | 323 |
| 79 | 284 |
| 75 | 265 |
| 51 | 4 |
| 50 | 2 |
| 67 | 2 |
| 52 | 1 |
| 68 | 1 |

**Table 11.**
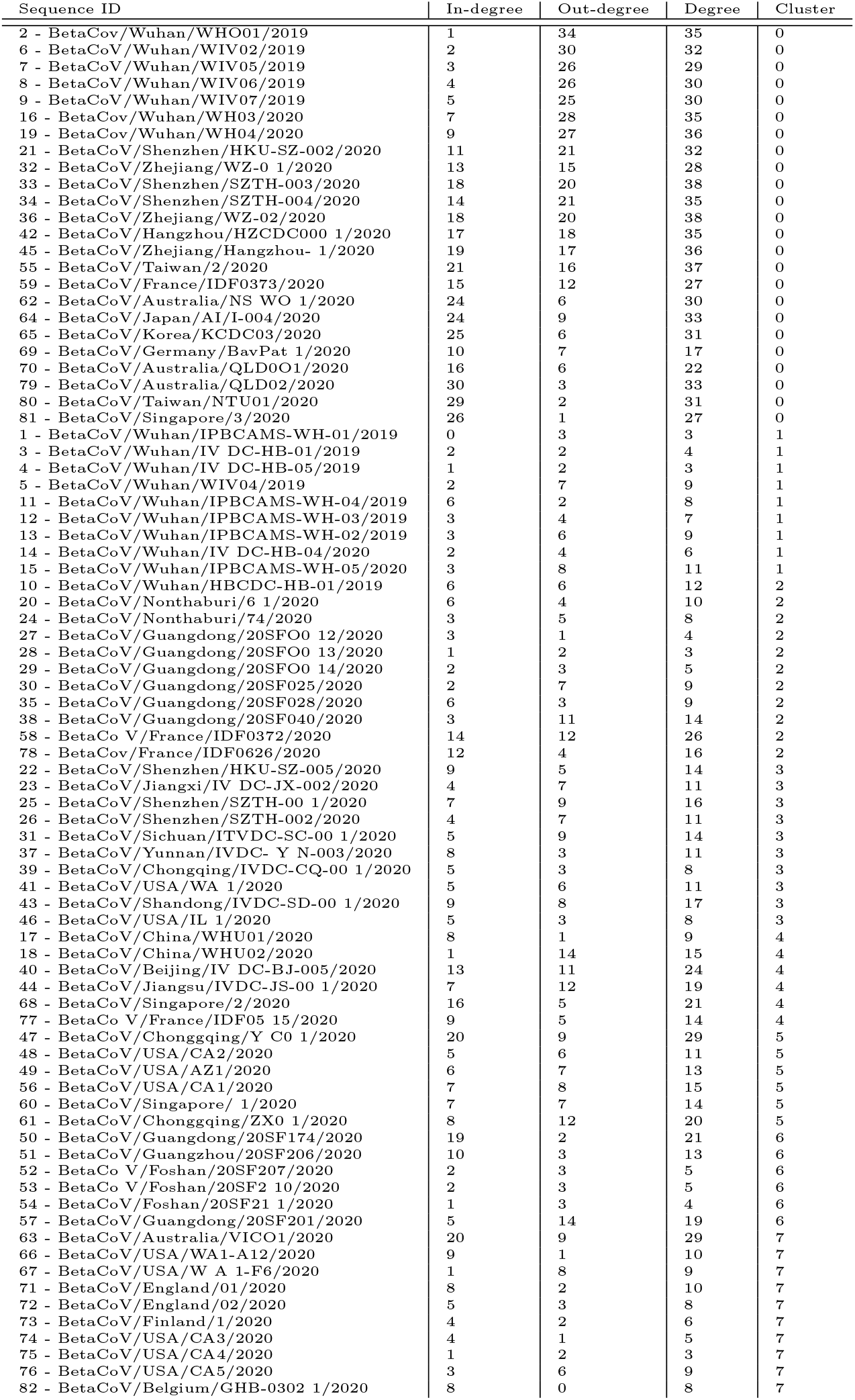
Cluster memberships of the nucleotide sequences (full TIC network).

- At the center of the network one cluster emerges (dark red, cluster 0) that has only few stronger connections within the cluster but features connections to all other clusters and within itself through lower weighted edges. This cluster has no temporal focus, it comprises samples that were collected early as well as later during the outbreak. While the genome sequences that represent the beginning of the progression stage path in this cluster are from China, it then transitions geographically to Australia, Japan, Korea, Germany, Taiwan and Singapore.
- The light red cluster (cluster 1) comprises a coherent sequence of samples taken from patients in the original outbreak region (Wuhan) and is temporally focused on the early progression stages.
- The orange cluster starting from the bottom center of the network (cluster 2) is again first a sequence of samples from the original outbreak region in Wuhan, transitioning to Thailand (Nonthaburi), then back to China (Guangdong), before finishing in France.
- Cluster 3 (yellow) features samples taken in various places in China but also two later connections to the USA.
- Cluster 4 (light green) is one of the smallest clusters but is spread fairly wide temporally. It features 4 samples taken in China and then transitions geographically to Singapore and France.
- In dark green (cluster 5) we can identify a cluster of genome data that was collected in Chonggqing, the USA and Singapore.
- The light blue cluster (cluster 6) is a temporally quite coherent sequence that links samples taken in Guangdong, Guangzhou and Foshan.
- A kind of international cluster (samples from Australia, the USA, England, Finland and Belgium) is represented by the dark blue cluster (cluster 7).

We suggest that while there are some obvious pathways of high similarity due to temporal order of the samples taken, there are still good indications for geographical-dependencies and transition phases that deserve further investigation. Transition points into and out of cluster 0 are particularly interesting in this regard, especially when the clusters linking in our out feature samples taken outside of China. We suggest that it may be worth to qualitatively assess the human-to-human connections the respective subjects had from which these samples were taken, and the places they have visited if there are no known links between the samples so far.

Triangulating the full TIC network with the networks that focus on individual reading frame similarity only (please refer to Figures 6, 7 and 8 as well as Tables 13, 15 and 17 in the appendix) shows that the macroscopic structure is mostly consistent but much sparser in terms of low-weighted edges.

**Table 12.** Inter-cluster similarities for the full TIC network.

| Inter-cluster pair | Avg. similarity | SD | Min. similarity | Max. similarity |
| --- | --- | --- | --- | --- |
| 0-1 | 0.267773 | 0.013962 | 0.252325 | 0.293298 |
| 0-2 | 0.265648 | 0.012247 | 0.251072 | 0.288222 |
| 0-3 | 0.267719 | 0.013965 | 0.252326 | 0.293298 |
| 0-4 | 0.267501 | 0.021119 | 0.250904 | 0.358635 |
| 0-5 | 0.267639 | 0.013011 | 0.252337 | 0.293264 |
| 0-6 | 0.265093 | 0.013211 | 0.251181 | 0.288327 |
| 0-7 | 0.267556 | 0.013985 | 0.250745 | 0.293466 |
| 1-2 | 0.259499 | 0.000047 | 0.259407 | 0.259605 |
| 1-3 | 0.999513 | 0.000290 | 0.998796 | 0.999900 |
| 1-4 | 0.269435 | 0.000147 | 0.269120 | 0.269655 |
| 1-5 | 0.927233 | 0.103628 | 0.781755 | 0.999966 |
| 1-6 | 0.258528 | 0.000033 | 0.258454 | 0.258598 |
| 1-7 | 0.998823 | 0.001846 | 0.993938 | 0.999933 |
| 2-3 | 0.259472 | 0.000053 | 0.259373 | 0.259572 |
| 2-4 | 0.255754 | 0.000079 | 0.255587 | 0.255887 |
| 2-5 | 0.260785 | 0.001855 | 0.259407 | 0.263405 |
| 2-6 | 0.284333 | 0.000043 | 0.284259 | 0.284454 |
| 2-7 | 0.259330 | 0.000421 | 0.258067 | 0.259682 |
| 3-4 | 0.269369 | 0.000184 | 0.268965 | 0.269823 |
| 3-5 | 0.926970 | 0.103499 | 0.781540 | 0.999967 |
| 3-6 | 0.258482 | 0.000049 | 0.258387 | 0.258598 |
| 3-7 | 0.998632 | 0.001816 | 0.993235 | 1.000000 |
| 4-5 | 0.268894 | 0.000682 | 0.267715 | 0.269603 |
| 4-6 | 0.263907 | 0.000187 | 0.263414 | 0.264057 |
| 4-7 | 0.269143 | 0.000600 | 0.267290 | 0.269928 |
| 5-6 | 0.260599 | 0.002943 | 0.258521 | 0.264821 |
| 5-7 | 0.926460 | 0.103206 | 0.779396 | 0.999900 |
| 6-7 | 0.258383 | 0.000525 | 0.256844 | 0.258889 |

**Table 13.**
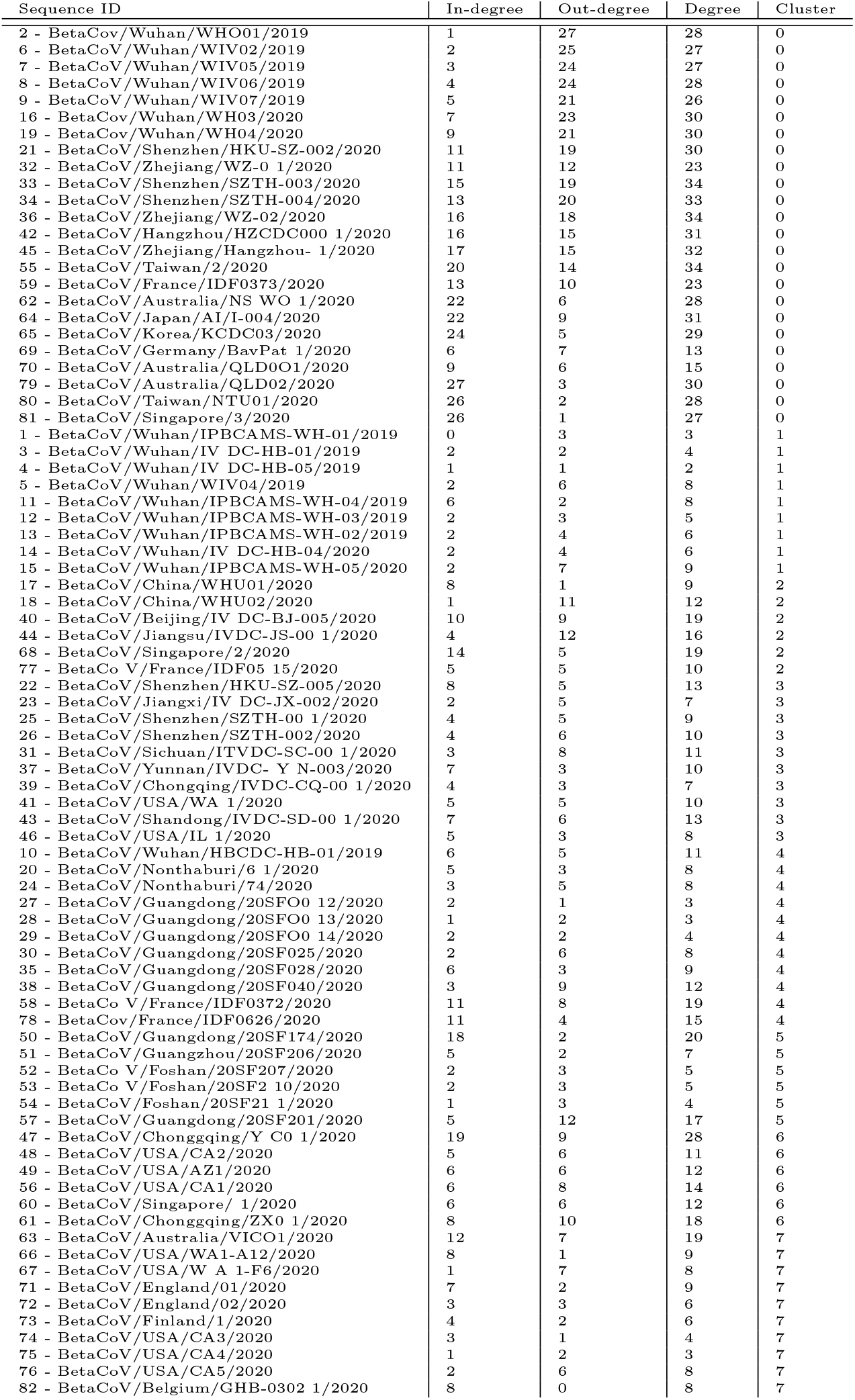
Cluster memberships of the nucleotide sequences (network for +1 reading frames).

| Sequence ID | In-degree | Out-degree | Degree | Cluster |
| --- | --- | --- | --- | --- |
| 2 - BetaCov/Wuhan/WHO01/2019 | 1 | 27 | 28 | 0 |
| 6 - BetaCoV/Wuhan/WIV02/2019 | 2 | 25 | 27 | 0 |
| 7 - BetaCoV/Wuhan/WIV05/2019 | 3 | 24 | 27 | 0 |
| 8 - BetaCoV/Wuhan/WIV06/2019 | 4 | 24 | 28 | 0 |
| 9 - BetaCoV/Wuhan/WIV07/2019 | 5 | 21 | 26 | 0 |
| 16 - BetaCov/Wuhan/WH03/2020 | 7 | 23 | 30 | 0 |
| 19 - BetaCov/Wuhan/WH04/2020 | 9 | 21 | 30 | 0 |
| 21 - BetaCoV/Shenzhen/HKU-SZ-002/2020 | 11 | 19 | 30 | 0 |
| 32 - BetaCoV/Zhejiang/WZ-0 1/2020 | 11 | 12 | 23 | 0 |
| 33 - BetaCoV/Shenzhen/SZTH-003/2020 | 15 | 19 | 34 | 0 |
| 34 - BetaCoV/Shenzhen/SZTH-004/2020 | 13 | 20 | 33 | 0 |
| 36 - BetaCoV/Zhejiang/WZ-02/2020 | 16 | 18 | 34 | 0 |
| 42 - BetaCoV/Hangzhou/HZCDC000 1/2020 | 16 | 15 | 31 | 0 |
| 45 - BetaCoV/Zhejiang/Hangzhou- 1/2020 | 17 | 15 | 32 | 0 |
| 55 - BetaCoV/Taiwan/2/2020 | 20 | 14 | 34 | 0 |
| 59 - BetaCoV/France/IDF0373/2020 | 13 | 10 | 23 | 0 |
| 62 - BetaCoV/Australia/NS WO 1/2020 | 22 | 6 | 28 | 0 |
| 64 - BetaCoV/Japan/AI/1-004/2020 | 22 | 9 | 31 | 0 |
| 65 - BetaCoV/Korea/KCDC03/2020 | 24 | 5 | 29 | 0 |
| 69 - BetaCoV/Germany/BavPat 1/2020 | 6 | 7 | 13 | 0 |
| 70 - BetaCoV/Australia/QLD001/2020 | 9 | 6 | 15 | 0 |
| 79 - BetaCoV/Australia/QLD02/2020 | 27 | 3 | 30 | 0 |
| 80 - BetaCoV/Taiwan/NTU01/2020 | 26 | 2 | 28 | 0 |
| 81 - BetaCoV/Singapore/3/2020 | 26 | 1 | 27 | 0 |
| 1 - BetaCoV/Wuhan/IPBCAMS-WH-01/2019 | 0 | 3 | 3 | 1 |
| 3 - BetaCoV/Wuhan/IV DC-HB-01/2019 | 2 | 2 | 4 | 1 |
| 4 - BetaCoV/Wuhan/IV DC-HB-05/2019 | 1 | 1 | 2 | 1 |
| 5 - BetaCoV/Wuhan/WIV04/2019 | 2 | 6 | 8 | 1 |
| 11 - BetaCoV/Wuhan/IPBCAMS-WH-04/2019 | 6 | 2 | 8 | 1 |
| 12 - BetaCoV/Wuhan/IPBCAMS-WH-03/2019 | 2 | 3 | 5 | 1 |
| 13 - BetaCoV/Wuhan/IPBCAMS-WH-02/2019 | 2 | 4 | 6 | 1 |
| 14 - BetaCoV/Wuhan/IV DC-HB-04/2020 | 2 | 4 | 6 | 1 |
| 15 - BetaCoV/Wuhan/IPBCAMS-WH-05/2020 | 2 | 7 | 9 | 1 |
| 17 - BetaCoV/China/WHU01/2020 | 8 | 1 | 9 | 2 |
| 18 - BetaCoV/China/WHU02/2020 | 1 | 11 | 12 | 2 |
| 40 - BetaCoV/Beijing/IV DC-BJ-005/2020 | 10 | 9 | 19 | 2 |
| 44 - BetaCoV/Jiangsu/IVDC-JS-00 1/2020 | 4 | 12 | 16 | 2 |
| 68 - BetaCoV/Singapore/2/2020 | 14 | 5 | 19 | 2 |
| 77 - BetaCo V/France/IDF05 15/2020 | 5 | 5 | 10 | 2 |
| 22 - BetaCoV/Shenzhen/HKU-SZ-005/2020 | 8 | 5 | 13 | 3 |
| 23 - BetaCoV/Jiangxi/IV DC-JX-002/2020 | 2 | 5 | 7 | 3 |
| 25 - BetaCoV/Shenzhen/SZTH-00 1/2020 | 4 | 5 | 9 | 3 |
| 26 - BetaCoV/Shenzhen/SZTH-002/2020 | 4 | 6 | 10 | 3 |
| 31 - BetaCoV/Sichuan/ITVDC-SC-00 1/2020 | 3 | 8 | 11 | 3 |
| 37 - BetaCoV/Yunnan/IVDC- Y N-003/2020 | 7 | 3 | 10 | 3 |
| 39 - BetaCoV/Chongqing/IVDC-CQ-00 1/2020 | 4 | 3 | 7 | 3 |
| 41 - BetaCoV/USA/WA 1/2020 | 5 | 5 | 10 | 3 |
| 43 - BetaCoV/Shandong/IVDC-SD-00 1/2020 | 7 | 6 | 13 | 3 |
| 46 - BetaCoV/USA/IL 1/2020 | 5 | 3 | 8 | 3 |
| 10 - BetaCoV/Wuhan/HBCDC-HB-01/2019 | 6 | 5 | 11 | 4 |
| 20 - BetaCoV/Nonhaburi/6 1/2020 | 5 | 3 | 8 | 4 |
| 24 - BetaCoV/Nonhaburi/74/2020 | 3 | 5 | 8 | 4 |
| 27 - BetaCoV/Guangdong/20SFO0 12/2020 | 2 | 1 | 3 | 4 |
| 28 - BetaCoV/Guangdong/20SFO0 13/2020 | 1 | 2 | 3 | 4 |
| 29 - BetaCoV/Guangdong/20SFO0 14/2020 | 2 | 2 | 4 | 4 |
| 30 - BetaCoV/Guangdong/20SF025/2020 | 2 | 6 | 8 | 4 |
| 35 - BetaCoV/Guangdong/20SF028/2020 | 6 | 3 | 9 | 4 |
| 38 - BetaCoV/Guangdong/20SF040/2020 | 3 | 9 | 12 | 4 |
| 58 - BetaCo V/France/IDF0372/2020 | 11 | 8 | 19 | 4 |
| 78 - BetaCov/France/IDF0626/2020 | 11 | 4 | 15 | 4 |
| 50 - BetaCoV/Guangdong/20SF174/2020 | 18 | 2 | 20 | 5 |
| 51 - BetaCoV/Guangzhou/20SF206/2020 | 5 | 2 | 7 | 5 |
| 52 - BetaCo V/Foshan/20SF207/2020 | 2 | 3 | 5 | 5 |
| 53 - BetaCo V/Foshan/20SF2 10/2020 | 2 | 3 | 5 | 5 |
| 54 - BetaCoV/Foshan/20SF21 1/2020 | 1 | 3 | 4 | 5 |
| 57 - BetaCoV/Guangdong/20SF201/2020 | 5 | 12 | 17 | 5 |
| 47 - BetaCoV/Chongqing/Y C0 1/2020 | 19 | 9 | 28 | 6 |
| 48 - BetaCoV/USA/CA2/2020 | 5 | 6 | 11 | 6 |
| 49 - BetaCoV/USA/AZ1/2020 | 6 | 6 | 12 | 6 |
| 56 - BetaCoV/USA/CA1/2020 | 6 | 8 | 14 | 6 |
| 60 - BetaCoV/Singapore/ 1/2020 | 6 | 6 | 12 | 6 |
| 61 - BetaCoV/Chongqing/ZX0 1/2020 | 8 | 10 | 18 | 6 |
| 63 - BetaCoV/Australia/VICO1/2020 | 12 | 7 | 19 | 7 |
| 66 - BetaCoV/USA/WA1-A12/2020 | 8 | 1 | 9 | 7 |
| 67 - BetaCoV/USA/W A 1-F6/2020 | 1 | 7 | 8 | 7 |
| 71 - BetaCoV/England/01/2020 | 7 | 2 | 9 | 7 |
| 72 - BetaCoV/England/02/2020 | 3 | 3 | 6 | 7 |
| 73 - BetaCoV/Finland/1/2020 | 4 | 2 | 6 | 7 |
| 74 - BetaCoV/USA/CA3/2020 | 3 | 1 | 4 | 7 |
| 75 - BetaCoV/USA/CA4/2020 | 1 | 2 | 3 | 7 |
| 76 - BetaCoV/USA/CA5/2020 | 2 | 6 | 8 | 7 |
| 82 - BetaCoV/Belgium/GHB-0302 1/2020 | 8 | 0 | 8 | 7 |

**Table 14.** Intra-cluster similarities (network for +1 reading frames).

| Cluster | Avg. similarity | SD | Min. similarity | Max. similarity | Cluster size |
| --- | --- | --- | --- | --- | --- |
| 0 | 0.277353 | 0.088247 | 0.250620 | 0.999866 | 24 |
| 1 | 0.999730 | 0.000222 | 0.999231 | 1.000000 | 9 |
| 2 | 0.998768 | 0.001345 | 0.996387 | 1.000000 | 6 |
| 3 | 0.999385 | 0.000371 | 0.998461 | 0.999933 | 10 |
| 4 | 0.999915 | 0.000048 | 0.999832 | 1.000000 | 11 |
| 5 | 0.999926 | 0.000036 | 0.999866 | 1.000000 | 6 |
| 6 | 0.883701 | 0.112398 | 0.781976 | 0.999899 | 6 |
| 7 | 0.997900 | 0.002419 | 0.991158 | 1.000000 | 10 |

**Table 15.** Cluster memberships of the nucleotide sequences (network for +2 reading frames).

| Sequence ID | In-degree | Out-degree | Degree | Cluster |
| --- | --- | --- | --- | --- |
| 2 - BetaCov/Wuhan/WHO01/2019 | 1 | 31 | 32 | 0 |
| 6 - BetaCoV/Wuhan/WIV02/2019 | 2 | 28 | 30 | 0 |
| 7 - BetaCoV/Wuhan/WIV05/2019 | 3 | 24 | 27 | 0 |
| 8 - BetaCoV/Wuhan/WIV06/2019 | 4 | 22 | 26 | 0 |
| 9 - BetaCoV/Wuhan/WIV07/2019 | 5 | 23 | 28 | 0 |
| 16 - BetaCov/Wuhan/WH03/2020 | 7 | 23 | 30 | 0 |
| 19 - BetaCov/Wuhan/WH04/2020 | 9 | 25 | 34 | 0 |
| 21 - BetaCoV/Shenzhen/HKU-SZ-002/2020 | 10 | 20 | 30 | 0 |
| 32 - BetaCoV/Zhejiang/WZ-0 1/2020 | 11 | 14 | 25 | 0 |
| 33 - BetaCoV/Shenzhen/SZTH-003/2020 | 16 | 19 | 35 | 0 |
| 34 - BetaCoV/Shenzhen/SZTH-004/2020 | 14 | 20 | 34 | 0 |
| 36 - BetaCoV/Zhejiang/WZ-02/2020 | 14 | 19 | 33 | 0 |
| 42 - BetaCoV/Hangzhou/HZCDC000 1/2020 | 16 | 18 | 34 | 0 |
| 45 - BetaCoV/Zhejiang/Hangzhou- 1/2020 | 16 | 16 | 32 | 0 |
| 55 - BetaCoV/Taiwan/2/2020 | 21 | 15 | 36 | 0 |
| 59 - BetaCoV/France/IDF0373/2020 | 14 | 12 | 26 | 0 |
| 62 - BetaCoV/Australia/NS WO 1/2020 | 22 | 5 | 27 | 0 |
| 64 - BetaCoV/Japan/AI/I-004/2020 | 22 | 8 | 30 | 0 |
| 65 - BetaCoV/Korea/KCDC03/2020 | 23 | 6 | 29 | 0 |
| 69 - BetaCoV/Germany/BavPat 1/2020 | 6 | 7 | 13 | 0 |
| 70 - BetaCoV/Australia/QLD001/2020 | 10 | 6 | 16 | 0 |
| 79 - BetaCoV/Australia/QLD02/2020 | 28 | 3 | 31 | 0 |
| 80 - BetaCoV/Taiwan/NTU01/2020 | 27 | 2 | 29 | 0 |
| 81 - BetaCoV/Singapore/3/2020 | 26 | 1 | 27 | 0 |
| 1 - BetaCoV/Wuhan/IPBCAMS-WH-01/2019 | 0 | 3 | 3 | 1 |
| 3 - BetaCoV/Wuhan/IV DC-HB-01/2019 | 2 | 2 | 4 | 1 |
| 4 - BetaCoV/Wuhan/IV DC-HB-05/2019 | 1 | 1 | 2 | 1 |
| 5 - BetaCoV/Wuhan/WIV04/2019 | 2 | 6 | 8 | 1 |
| 11 - BetaCoV/Wuhan/IPBCAMS-WH-04/2019 | 6 | 2 | 8 | 1 |
| 12 - BetaCoV/Wuhan/IPBCAMS-WH-03/2019 | 2 | 3 | 5 | 1 |
| 13 - BetaCoV/Wuhan/IPBCAMS-WH-02/2019 | 2 | 3 | 5 | 1 |
| 14 - BetaCoV/Wuhan/IV DC-HB-04/2020 | 2 | 3 | 5 | 1 |
| 15 - BetaCoV/Wuhan/IPBCAMS-WH-05/2020 | 2 | 7 | 9 | 1 |
| 17 - BetaCoV/China/WHU01/2020 | 8 | 1 | 9 | 2 |
| 18 - BetaCoV/China/WHU02/2020 | 1 | 13 | 14 | 2 |
| 40 - BetaCoV/Beijing/IV DC-BJ-005/2020 | 12 | 9 | 21 | 2 |
| 44 - BetaCoV/Jiangsu/IVDC-JS-00 1/2020 | 6 | 11 | 17 | 2 |
| 68 - BetaCoV/Singapore/2/2020 | 14 | 4 | 18 | 2 |
| 77 - BetaCo V/France/IDF05 15/2020 | 7 | 5 | 12 | 2 |
| 10 - BetaCoV/Wuhan/HBCDC-HB-01/2019 | 6 | 6 | 12 | 3 |
| 20 - BetaCoV/Nonthaburi/6 1/2020 | 6 | 4 | 10 | 3 |
| 24 - BetaCoV/Nonthaburi/74/2020 | 3 | 4 | 7 | 3 |
| 27 - BetaCoV/Guangdong/20SFO0 12/2020 | 3 | 1 | 4 | 3 |
| 28 - BetaCoV/Guangdong/20SFO0 13/2020 | 1 | 2 | 3 | 3 |
| 29 - BetaCoV/Guangdong/20SFO0 14/2020 | 2 | 3 | 5 | 3 |
| 30 - BetaCoV/Guangdong/20SF025/2020 | 2 | 6 | 8 | 3 |
| 35 - BetaCoV/Guangdong/20SF028/2020 | 6 | 3 | 9 | 3 |
| 38 - BetaCoV/Guangdong/20SF040/2020 | 3 | 10 | 13 | 3 |
| 58 - BetaCo V/France/IDF0372/2020 | 12 | 9 | 21 | 3 |
| 78 - BetaCov/France/IDF0626/2020 | 11 | 4 | 15 | 3 |
| 22 - BetaCoV/Shenzhen/HKU-SZ-005/2020 | 8 | 4 | 12 | 4 |
| 23 - BetaCoV/Jiangxi/IV DC-JX-002/2020 | 4 | 5 | 9 | 4 |
| 25 - BetaCoV/Shenzhen/SZTH-00 1/2020 | 7 | 5 | 12 | 4 |
| 26 - BetaCoV/Shenzhen/SZTH-002/2020 | 3 | 6 | 9 | 4 |
| 31 - BetaCoV/Sichuan/ITVDC-SC-00 1/2020 | 4 | 9 | 13 | 4 |
| 37 - BetaCoV/Yunnan/IVDC- Y N-003/2020 | 8 | 3 | 11 | 4 |
| 39 - BetaCoV/Chongqing/IVDC-CQ-00 1/2020 | 4 | 3 | 7 | 4 |
| 41 - BetaCoV/USA/WA 1/2020 | 5 | 5 | 10 | 4 |
| 43 - BetaCoV/Shandong/IVDC-SD-00 1/2020 | 6 | 7 | 13 | 4 |
| 46 - BetaCoV/USA/IL 1/2020 | 5 | 3 | 8 | 4 |
| 50 - BetaCoV/Guangdong/20SF174/2020 | 19 | 2 | 21 | 5 |
| 51 - BetaCoV/Guangzhou/20SF206/2020 | 7 | 3 | 10 | 5 |
| 52 - BetaCo V/Foshan/20SF207/2020 | 2 | 2 | 4 | 5 |
| 53 - BetaCo V/Foshan/20SF2 10/2020 | 2 | 2 | 4 | 5 |
| 54 - BetaCoV/Foshan/20SF21 1/2020 | 1 | 3 | 4 | 5 |
| 57 - BetaCoV/Guangdong/20SF201/2020 | 5 | 12 | 17 | 5 |
| 47 - BetaCoV/Chongqing/Y C0 1/2020 | 20 | 9 | 29 | 6 |
| 48 - BetaCoV/USA/CA2/2020 | 4 | 4 | 8 | 6 |
| 49 - BetaCoV/USA/AZ1/2020 | 6 | 5 | 11 | 6 |
| 56 - BetaCoV/USA/CA1/2020 | 6 | 7 | 13 | 6 |
| 60 - BetaCoV/Singapore/ 1/2020 | 6 | 5 | 11 | 6 |
| 61 - BetaCoV/Chongqing/ZX0 1/2020 | 7 | 10 | 17 | 6 |
| 63 - BetaCoV/Australia/VICO1/2020 | 15 | 7 | 22 | 7 |
| 66 - BetaCoV/USA/WA1-A12/2020 | 7 | 1 | 8 | 7 |
| 67 - BetaCoV/USA/W A 1-F6/2020 | 1 | 8 | 9 | 7 |
| 71 - BetaCoV/England/01/2020 | 6 | 2 | 8 | 7 |
| 72 - BetaCoV/England/02/2020 | 2 | 3 | 5 | 7 |
| 73 - BetaCoV/Finland/1/2020 | 4 | 2 | 6 | 7 |
| 74 - BetaCoV/USA/CA3/2020 | 4 | 1 | 5 | 7 |
| 75 - BetaCoV/USA/CA4/2020 | 1 | 2 | 3 | 7 |
| 76 - BetaCoV/USA/CA5/2020 | 2 | 6 | 8 | 7 |
| 82 - BetaCoV/Belgium/GHB-0302 1/2020 | 8 | 0 | 8 | 7 |

**Table 16.** Intra-cluster similarities (network for +2 reading frames).

| Cluster | Avg. similarity | SD | Min. similarity | Max. similarity | Cluster size |
| --- | --- | --- | --- | --- | --- |
| 0 | 0.277353 | 0.088247 | 0.250620 | 0.999866 | 24 |
| 1 | 0.999730 | 0.000222 | 0.999231 | 1.000000 | 9 |
| 2 | 0.998768 | 0.001345 | 0.996387 | 1.000000 | 6 |
| 3 | 0.999915 | 0.000048 | 0.999832 | 1.000000 | 11 |
| 4 | 0.999385 | 0.000371 | 0.998461 | 0.999933 | 10 |
| 5 | 0.999926 | 0.000036 | 0.999866 | 1.000000 | 6 |
| 6 | 0.883701 | 0.112398 | 0.781976 | 0.999899 | 6 |
| 7 | 0.997900 | 0.002419 | 0.991158 | 1.000000 | 10 |

**Table 17.**
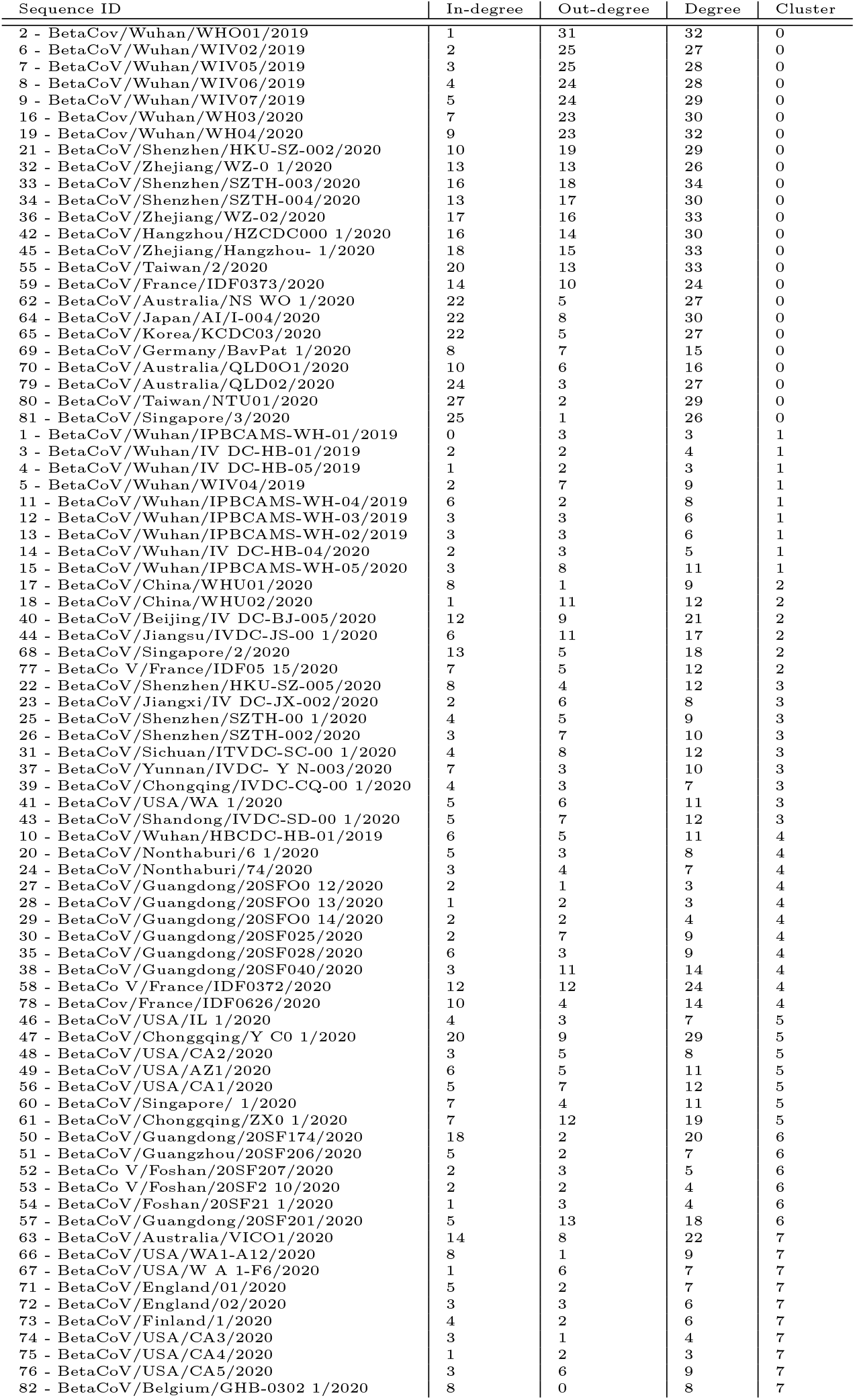
Cluster memberships of the nucleotide sequences (network for +3 reading frames).

| Sequence ID | In-degree | Out-degree | Degree | Cluster |
| --- | --- | --- | --- | --- |
| 2 - BetaCov/Wuhan/WHO01/2019 | 1 | 31 | 32 | 0 |
| 6 - BetaCoV/Wuhan/WIV02/2019 | 2 | 25 | 27 | 0 |
| 7 - BetaCoV/Wuhan/WIV05/2019 | 3 | 25 | 28 | 0 |
| 8 - BetaCoV/Wuhan/WIV06/2019 | 4 | 24 | 28 | 0 |
| 9 - BetaCoV/Wuhan/WIV07/2019 | 5 | 24 | 29 | 0 |
| 16 - BetaCov/Wuhan/WH03/2020 | 7 | 23 | 30 | 0 |
| 19 - BetaCov/Wuhan/WH04/2020 | 9 | 23 | 32 | 0 |
| 21 - BetaCoV/Shenzhen/HKU-SZ-002/2020 | 10 | 19 | 29 | 0 |
| 32 - BetaCoV/Zhejiang/WZ-0 1/2020 | 13 | 13 | 26 | 0 |
| 33 - BetaCoV/Shenzhen/SZTH-003/2020 | 16 | 18 | 34 | 0 |
| 34 - BetaCoV/Shenzhen/SZTH-004/2020 | 13 | 17 | 30 | 0 |
| 36 - BetaCoV/Zhejiang/WZ-02/2020 | 17 | 16 | 33 | 0 |
| 42 - BetaCoV/Hangzhou/HZCDC000 1/2020 | 16 | 14 | 30 | 0 |
| 45 - BetaCoV/Zhejiang/Hangzhou- 1/2020 | 18 | 15 | 33 | 0 |
| 55 - BetaCoV/Taiwan/2/2020 | 20 | 13 | 33 | 0 |
| 59 - BetaCoV/France/IDF0373/2020 | 14 | 10 | 24 | 0 |
| 62 - BetaCoV/Australia/NS WO 1/2020 | 22 | 5 | 27 | 0 |
| 64 - BetaCoV/Japan/AI/1-004/2020 | 22 | 8 | 30 | 0 |
| 65 - BetaCoV/Korea/KCDC03/2020 | 22 | 5 | 27 | 0 |
| 69 - BetaCoV/Germany/BavPat 1/2020 | 8 | 7 | 15 | 0 |
| 70 - BetaCoV/Australia/QLD001/2020 | 10 | 6 | 16 | 0 |
| 79 - BetaCoV/Australia/QLD02/2020 | 24 | 3 | 27 | 0 |
| 80 - BetaCoV/Taiwan/NTU01/2020 | 27 | 2 | 29 | 0 |
| 81 - BetaCoV/Singapore/3/2020 | 25 | 1 | 26 | 0 |
| 1 - BetaCoV/Wuhan/IPBCAMS-WH-01/2019 | 0 | 3 | 3 | 1 |
| 3 - BetaCoV/Wuhan/IV DC-HB-01/2019 | 2 | 2 | 4 | 1 |
| 4 - BetaCoV/Wuhan/IV DC-HB-05/2019 | 1 | 2 | 3 | 1 |
| 5 - BetaCoV/Wuhan/WIV04/2019 | 2 | 7 | 9 | 1 |
| 11 - BetaCoV/Wuhan/IPBCAMS-WH-04/2019 | 6 | 2 | 8 | 1 |
| 12 - BetaCoV/Wuhan/IPBCAMS-WH-03/2019 | 3 | 3 | 6 | 1 |
| 13 - BetaCoV/Wuhan/IPBCAMS-WH-02/2019 | 3 | 3 | 6 | 1 |
| 14 - BetaCoV/Wuhan/IV DC-HB-04/2020 | 2 | 3 | 5 | 1 |
| 15 - BetaCoV/Wuhan/IPBCAMS-WH-05/2020 | 3 | 8 | 11 | 1 |
| 17 - BetaCoV/China/WHU01/2020 | 8 | 1 | 9 | 2 |
| 18 - BetaCoV/China/WHU02/2020 | 1 | 11 | 12 | 2 |
| 40 - BetaCoV/Beijing/IV DC-BJ-005/2020 | 12 | 9 | 21 | 2 |
| 44 - BetaCoV/Jiangsu/IVDC-JS-00 1/2020 | 6 | 11 | 17 | 2 |
| 68 - BetaCoV/Singapore/2/2020 | 13 | 5 | 18 | 2 |
| 77 - BetaCo V/France/IDF05 15/2020 | 7 | 5 | 12 | 2 |
| 22 - BetaCoV/Shenzhen/HKU-SZ-005/2020 | 8 | 4 | 12 | 3 |
| 23 - BetaCoV/Jiangxi/IV DC-JX-002/2020 | 2 | 6 | 8 | 3 |
| 25 - BetaCoV/Shenzhen/SZTH-00 1/2020 | 4 | 5 | 9 | 3 |
| 26 - BetaCoV/Shenzhen/SZTH-002/2020 | 3 | 7 | 10 | 3 |
| 31 - BetaCoV/Sichuan/ITVDC-SC-00 1/2020 | 4 | 8 | 12 | 3 |
| 37 - BetaCoV/Yunnan/IVDC- Y N-003/2020 | 7 | 3 | 10 | 3 |
| 39 - BetaCoV/Chongqing/IVDC-CQ-00 1/2020 | 4 | 3 | 7 | 3 |
| 41 - BetaCoV/USA/WA 1/2020 | 5 | 6 | 11 | 3 |
| 43 - BetaCoV/Shandong/IVDC-SD-00 1/2020 | 5 | 7 | 12 | 3 |
| 10 - BetaCoV/Wuhan/HBCDC-HB-01/2019 | 6 | 5 | 11 | 4 |
| 20 - BetaCoV/Nonthaburi/6 1/2020 | 5 | 3 | 8 | 4 |
| 24 - BetaCoV/Nonthaburi/74/2020 | 3 | 4 | 7 | 4 |
| 27 - BetaCoV/Guangdong/20SFO0 12/2020 | 2 | 1 | 3 | 4 |
| 28 - BetaCoV/Guangdong/20SFO0 13/2020 | 1 | 2 | 3 | 4 |
| 29 - BetaCoV/Guangdong/20SFO0 14/2020 | 2 | 2 | 4 | 4 |
| 30 - BetaCoV/Guangdong/20SF025/2020 | 2 | 7 | 9 | 4 |
| 35 - BetaCoV/Guangdong/20SF028/2020 | 6 | 3 | 9 | 4 |
| 38 - BetaCoV/Guangdong/20SF040/2020 | 3 | 11 | 14 | 4 |
| 58 - BetaCo V/France/IDF0372/2020 | 12 | 12 | 24 | 4 |
| 78 - BetaCov/France/IDF0626/2020 | 10 | 4 | 14 | 4 |
| 46 - BetaCoV/USA/IL 1/2020 | 4 | 3 | 7 | 5 |
| 47 - BetaCoV/Chongqing/Y C0 1/2020 | 20 | 9 | 29 | 5 |
| 48 - BetaCoV/USA/CA2/2020 | 3 | 5 | 8 | 5 |
| 49 - BetaCoV/USA/AZ1/2020 | 6 | 5 | 11 | 5 |
| 56 - BetaCoV/USA/CA1/2020 | 5 | 7 | 12 | 5 |
| 60 - BetaCoV/Singapore/ 1/2020 | 7 | 4 | 11 | 5 |
| 61 - BetaCoV/Chongqing/ZX0 1/2020 | 7 | 12 | 19 | 5 |
| 50 - BetaCoV/Guangdong/20SF174/2020 | 18 | 2 | 20 | 6 |
| 51 - BetaCoV/Guangzhou/20SF206/2020 | 5 | 2 | 7 | 6 |
| 52 - BetaCo V/Foshan/20SF207/2020 | 2 | 3 | 5 | 6 |
| 53 - BetaCo V/Foshan/20SF2 10/2020 | 2 | 2 | 4 | 6 |
| 54 - BetaCoV/Foshan/20SF21 1/2020 | 1 | 3 | 4 | 6 |
| 57 - BetaCoV/Guangdong/20SF201/2020 | 5 | 13 | 18 | 6 |
| 63 - BetaCoV/Australia/VICO1/2020 | 14 | 8 | 22 | 7 |
| 66 - BetaCoV/USA/WA1-A12/2020 | 8 | 1 | 9 | 7 |
| 67 - BetaCoV/USA/W A 1-F6/2020 | 1 | 6 | 7 | 7 |
| 71 - BetaCoV/England/01/2020 | 5 | 2 | 7 | 7 |
| 72 - BetaCoV/England/02/2020 | 3 | 3 | 6 | 7 |
| 73 - BetaCoV/Finland/1/2020 | 4 | 2 | 6 | 7 |
| 74 - BetaCoV/USA/CA3/2020 | 3 | 1 | 4 | 7 |
| 75 - BetaCoV/USA/CA4/2020 | 1 | 2 | 3 | 7 |
| 76 - BetaCoV/USA/CA5/2020 | 3 | 6 | 9 | 7 |
| 82 - BetaCoV/Belgium/GHB-0302 1/2020 | 8 | 0 | 8 | 7 |

**Figure 8.**
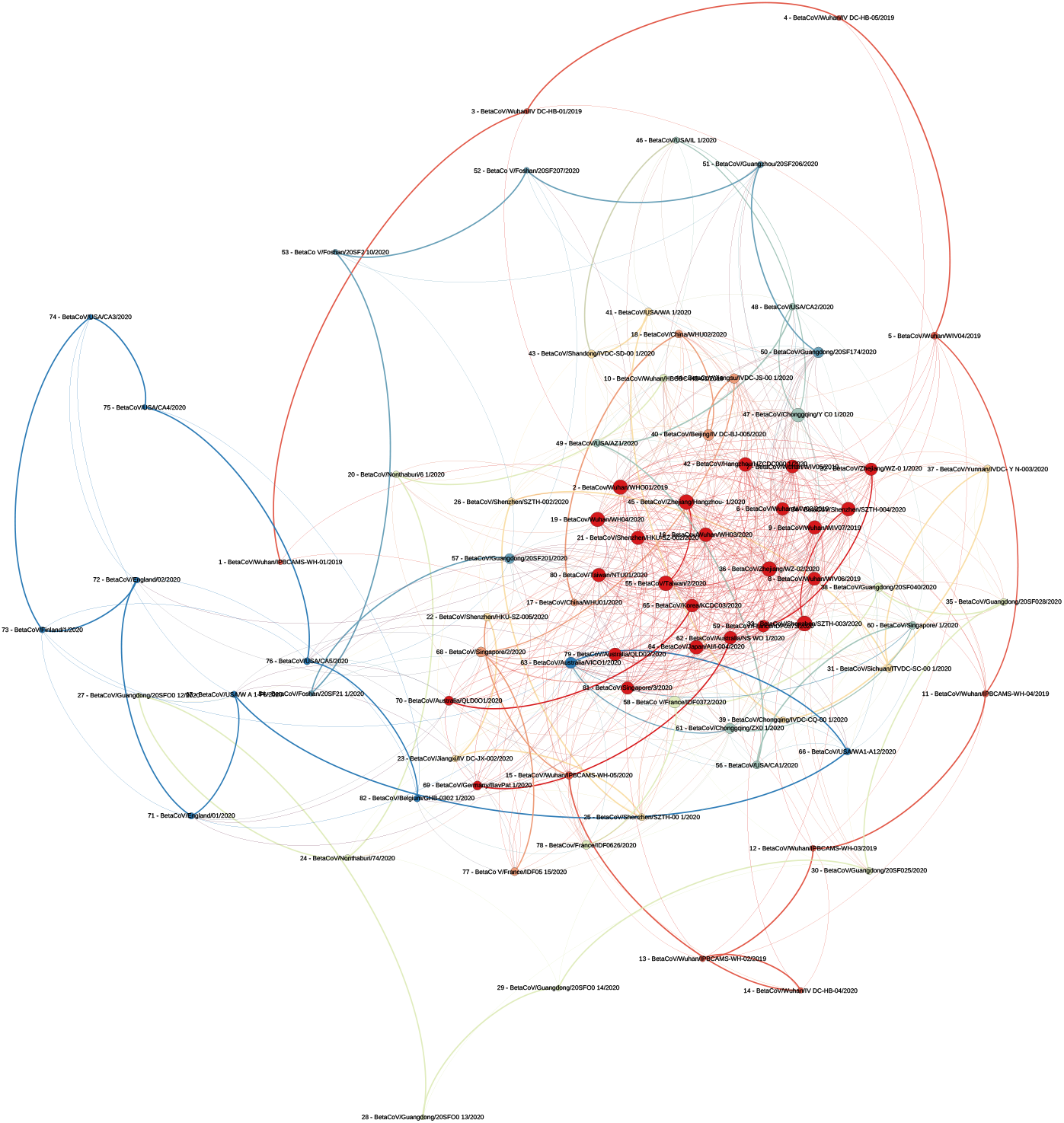
+3 reading frame network.

### Inter- and intra-cluster similarity

From our measurement of the intra-cluster similarity (see Table 1) we find that the genomic similarity at the level of nucleotides is low in the central cluster 0, which is expected because this is the largest of all clusters with low temporal coherence. However, cluster 5 also features a slightly lower average similarity with comparably high standard deviation, which indicates that this cluster may be slightly less coherent.

The barchart of the inter-cluster analysis results shown in Figure 5 reveals that there are some cluster pairs with high inter-cluster similarity, that the inter-cluster similarity is different between the +3 reading frame network and the other two reading frame networks, and that the pattern of the +3 reading frame network inter-cluster similarity is similar to the one for the entire TIC network. In particular we find that in the +1 and +2 reading frame networks the clusters 1, 4, 6 and 7 are quite similar at the nucleotide level compared to all other clusters that are pairwise distinct. For the entire TIC network and the +3 reading frame network instead we find that clusters 1, 3, 5 and 7 show this kind of similarity. Altogether, these results indicate that (a) in some instances the clustering based on structural characteristics of the TIC network overrides actual genomic similarity, and (b) the choice of the reading frames during TIC construction has an impact on the resulting network that needs to be further investigated.

## Conclusion

In this paper we presented a novel way to model, visualise and analyse the temporal dynamics of an epidemic outbreak. We applied Transcendental Information Cascades to capture the recurrence and co-occurrence of unique codon identifiers over distinct and temporally ordered nucleotide sequences derived from genetic samples taken from different human subjects. Our case study was focused on the recent outbreak of the novel 2019-nCoV coronavirus. We find clusters and evolution pathways of virus genomes in the constructed Transcendental Information Cascade that are candidates to be investigated further for potential human-to-human transmission paths and genetic evolution of the 2019-nCoV coronavirus.

Our study was performed as a snapshot case study on a limited sample of virus data. While this was at the time of this study all data that was available, we will need to re-execute this analysis continuously as more data becomes available to check whether any of our results change. Furthermore, our results indicate that experiments with alternative tokenisations are needed, such as weighting the +1, +2 and +3 reading frames differently (or omitting certain frames entirely) depending on their functional role [10]. We also suggest that it will be important to perform a wider variety of analysis of the generated TIC data (e.g. testing of different network clustering methods, application of dynamical systems analyses techniques) to get more profound insights. Our analysis also does not take measurement biases into account that may results from different techniques used by different laboratories for genome extraction and analysis [7].

In summary, the approach presented in this paper is exploratory and hypotheses generating in nature. It does not confirm or reject any of the hypotheses we generated about the potential transmission and evolution pathways nor does it provide causal explanation for the patterns found. However, the purpose of our work is to provide an alternative view to the epidemic dynamics of the 2019-nCoV outbreak (and potential other outbreaks in the future) that is hidden to state-of-the-art methods, and that may trigger geneticists and epidemiologists to look at pathways in epidemic outbreaks they would not normally be aware of.

## Data Availability

This study uses secondary data that was obtained from the GISAID EpiFlu™ Database. The software used in this study is available as open source software to allow for reproduction of our analysis.

https://www.gisaid.org/

https://github.com/vuw-c2lab/transcendental-information-cascades/tree/master/input/2020-02-03-2019nCoV

## Appendix

The software developed for this particular analysis is part of the open scientific software “R toolchain to construct and analyse Transcendental Information Cascades” that is available via https://github.com/vuw-c2lab/transcendental-information-cascades/tree/master/input/2020-02-03-2019nCoV. For reproduction purposes it is necessary to obtain the raw genomic data used in this study from the GISAID EpiFlu Database™. For further reference we provide the raw results of some of our analysis. We particularly point to the tables that provide references for relationships between nucleotide sequences and the cluster membership of nucleotide sequences.

## Acknowledgements

We want to thank the laboratories and researchers who contributed the genomic research data used in this study and obtained via the *GISAID EpiFlu Database*™ ^3^ on Monday, February 10th 2020. The following tables contain information about the origination of the research data used in this study and give credit to the collecting and submitting researchers and laboratories.

---

**Table 18.**
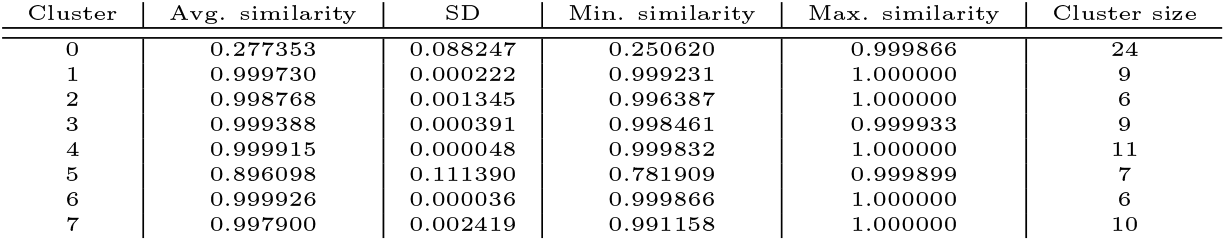
Intra-cluster similarities (network for +3 reading frames).

| Cluster | Avg. similarity | SD | Min. similarity | Max. similarity | Cluster size |
| --- | --- | --- | --- | --- | --- |
| 0 | 0.277353 | 0.088247 | 0.250620 | 0.999866 | 24 |
| 1 | 0.999730 | 0.000222 | 0.999231 | 1.000000 | 9 |
| 2 | 0.998768 | 0.001345 | 0.996387 | 1.000000 | 6 |
| 3 | 0.999388 | 0.000391 | 0.998461 | 0.999933 | 9 |
| 4 | 0.999915 | 0.000048 | 0.999832 | 1.000000 | 11 |
| 5 | 0.896098 | 0.111390 | 0.781909 | 0.999899 | 7 |
| 6 | 0.999926 | 0.000036 | 0.999866 | 1.000000 | 6 |
| 7 | 0.997900 | 0.002419 | 0.991158 | 1.000000 | 10 |

**Table 19.**
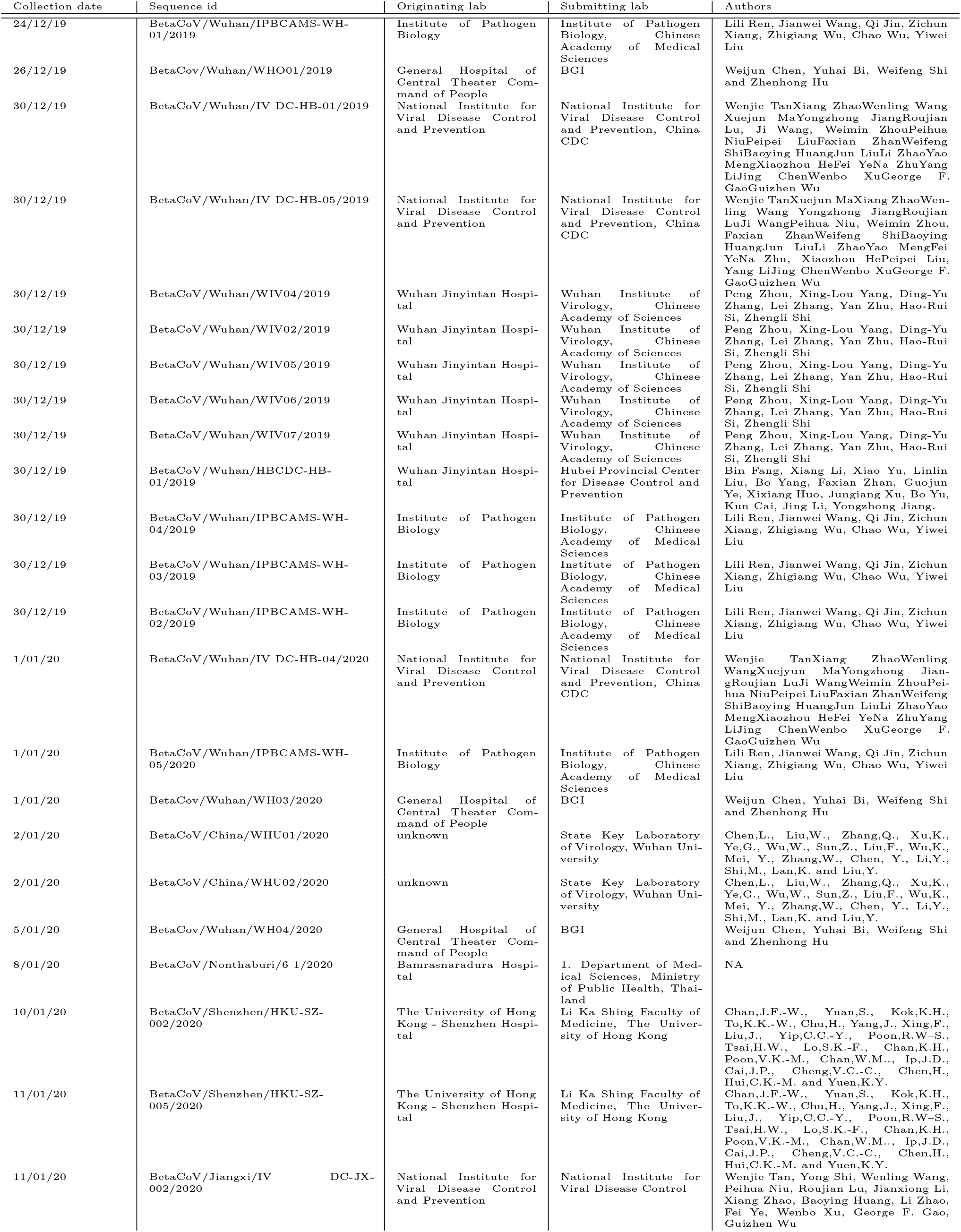
Nucleotide sequences used in this study that were obtained from the GISAID EpiFlu Database™.

**Table 20.** Nucleotide sequences used in this study that were obtained from the GISAID EpiFlu Database™(contd.).

| Collection date | Sequence id | Originating lab | Submitting lab | Authors |
| --- | --- | --- | --- | --- |
| 13/01/20 | BetaCoV/Nonthaburi/74/2020 | Bamrasnaradura Hospi-<br>tal | 1. Department of Med-<br>ical Sciences, Ministry<br>of Public Health, Thai-<br>land | NA |
| 13/01/20 | BetaCoV/Shenzhen/SZTH-00 1/2020 | Shenzhen Third People | Shenzhen Key Labo-<br>ratory of Pathogen<br>and Immunity, Na-<br>tional Clinical Research<br>Center for Infectious<br>Disease, | Yang Yang, Chenguang Shen, Li Xing,<br>Zhixiang Xu, Haixia Zheng, Yingxia<br>Liu |
| 13/01/20 | BetaCoV/Shenzhen/SZTH-002/2020 | Shenzhen Key Labora-<br>tory of Pathogen and<br>Immunity | Shenzhen Key Labo-<br>ratory of Pathogen<br>and Immunity, Na-<br>tional Clinical Research<br>Center for Infectious<br>Disease, | Yang Yang, Chenguang Shen, Li Xing,<br>Zhixiang Xu, Haixia Zheng, Yingxia<br>Liu |
| 14/01/20 | BetaCoV/Guangdong/20SFO0<br>12/2020 | Guangdong Provincial<br>Center for Diseases<br>Control and Prevention | Department of Micro-<br>biology, Guangdong<br>Provincial Center for<br>Diseases Control and<br>Prevention | Min Kang, Jie Wu, Jing Lu, Tao<br>Liu, Baisheng Li, Shuijiang Mei, Feng<br>Ruan, Lifeng Lin, Changwen Ke, Hao-<br>jie Zhong, Yingtao Zhang, Lirong<br>Zou, Xuguang Chen, Qi Zhu, Jian-<br>peng Xiao, Jianxiang Geng, Zhe Liu,<br>Jianxiang Hu, Weilin Zeng, Xing<br>Li, Yuhuang Liao, Xiujuan Tang,<br>Songjian Xiao, Ying Wang, Yingchao<br>Song, Xue Zhuang, Liyun Liang,<br>Guanhao He, Huihong Deng, Tie Song,<br>Jianfeng He, Wenjun Ma |
| 15/01/20 | BetaCoV/Guangdong/20SFO0<br>13/2020 | Guangdong Provincial<br>Center for Diseases<br>Control and Prevention | Department of Micro-<br>biology, Guangdong<br>Provincial Center for<br>Diseases Control and<br>Prevention | Min Kang, Jie Wu, Jing Lu, Tao<br>Liu, Baisheng Li, Shuijiang Mei, Feng<br>Ruan, Lifeng Lin, Changwen Ke, Hao-<br>jie Zhong, Yingtao Zhang, Lirong<br>Zou, Xuguang Chen, Qi Zhu, Jian-<br>peng Xiao, Jianxiang Geng, Zhe Liu,<br>Jianxiang Hu, Weilin Zeng, Xing<br>Li, Yuhuang Liao, Xiujuan Tang,<br>Songjian Xiao, Ying Wang, Yingchao<br>Song, Xue Zhuang, Liyun Liang,<br>Guanhao He, Huihong Deng, Tie Song,<br>Jianfeng He, Wenjun Ma |
| 15/01/20 | BetaCoV/Guangdong/20SFO0<br>14/2020 | Guangdong Provincial<br>Center for Diseases<br>Control and Prevention | Department of Micro-<br>biology, Guangdong<br>Provincial Center for<br>Diseases Control and<br>Prevention | Min Kang, Jie Wu, Jing Lu, Tao<br>Liu, Baisheng Li, Shuijiang Mei, Feng<br>Ruan, Lifeng Lin, Changwen Ke, Hao-<br>jie Zhong, Yingtao Zhang, Lirong<br>Zou, Xuguang Chen, Qi Zhu, Jian-<br>peng Xiao, Jianxiang Geng, Zhe Liu,<br>Jianxiang Hu, Weilin Zeng, Xing<br>Li, Yuhuang Liao, Xiujuan Tang,<br>Songjian Xiao, Ying Wang, Yingchao<br>Song, Xue Zhuang, Liyun Liang,<br>Guanhao He, Huihong Deng, Tie Song,<br>Jianfeng He, Wenjun Ma |
| 15/01/20 | BetaCoV/Guangdong/20SF025/2020 | Guangdong Provincial<br>Center for Diseases<br>Control and Prevention | Department of Micro-<br>biology, Guangdong<br>Provincial Center for<br>Diseases Control and<br>Prevention | Min Kang, Jie Wu, Jing Lu, Tao<br>Liu, Baisheng Li, Shuijiang Mei, Feng<br>Ruan, Lifeng Lin, Changwen Ke, Hao-<br>jie Zhong, Yingtao Zhang, Lirong<br>Zou, Xuguang Chen, Qi Zhu, Jian-<br>peng Xiao, Jianxiang Geng, Zhe Liu,<br>Jianxiang Hu, Weilin Zeng, Xing<br>Li, Yuhuang Liao, Xiujuan Tang,<br>Songjian Xiao, Ying Wang, Yingchao<br>Song, Xue Zhuang, Liyun Liang,<br>Guanhao He, Huihong Deng, Tie Song,<br>Jianfeng He, Wenjun Ma |
| 15/01/20 | BetaCoV/Sichuan/ITVDC-SC-00<br>1/2020 | National Institute for<br>Viral Disease Control<br>and Prevention | National Institute for<br>Viral Disease Control<br>and Prevention | Wenjie Tan, Jianan Xu, Wenling<br>Wang, Peihua Niu, Roujian Lu, Huip-<br>ing Yang, Xiang Zhao, Baoying Huang,<br>Li Zhao, Fei Ye, Wenbo Xu, George F.<br>Gao, Guizhen Wu |
| 16/01/20 | BetaCoV/Zhejiang/WZ-0 1/2020 | Zhejiang Provincial<br>Center for Disease<br>Control and Prevention | Department of Microbi-<br>ology, Zhejiang Provin-<br>cial Center for Disease<br>Control and Prevention | Yin Chen, Yanjun Zhang, Haiyan Mao,<br>Junhang Pan, Xiuyu Lou, Yiyu Lu,<br>Juying Yan, Hanping Zhu, Jian Gao,<br>Yan Feng, Yi Sun, Hao Yan, Zhen Li,<br>Yisheng Sun, Liming Gong, Qiong Ge,<br>Wen Shi, Xinying Wang, Wenwu Yao,<br>Zhangnv Yang, Fang Xu, Chen Chen,<br>Enfu Chen, Zhen Wang, Zhiping Chen,<br>Jianmin Jiang, Chonggao Hu |
| 16/01/20 | BetaCoV/Shenzhen/SZTH-003/2020 | Shenzhen Key Labora-<br>tory of Pathogen and<br>Immunity | Shenzhen Key Labo-<br>ratory of Pathogen<br>and Immunity, Na-<br>tional Clinical Research<br>Center for Infectious<br>Disease, | Yang Yang, Chenguang Shen, Li Xing,<br>Zhixiang Xu, Haixia Zheng, Yingxia<br>Liu |
| 16/01/20 | BetaCoV/Shenzhen/SZTH-004/2020 | Shenzhen Key Labora-<br>tory of Pathogen and<br>Immunity | Shenzhen Key Labo-<br>ratory of Pathogen<br>and Immunity, Na-<br>tional Clinical Research<br>Center for Infectious<br>Disease, | Yang Yang, Chenguang Shen, Li Xing,<br>Zhixiang Xu, Haixia Zheng, Yingxia<br>Liu |
| 17/01/20 | BetaCoV/Guangdong/20SF028/2020 | Guangdong Provincial<br>Center for Diseases<br>Control and Prevention | Department of Micro-<br>biology, Guangdong<br>Provincial Center for<br>Diseases Control and<br>Prevention | Min Kang, Jie Wu, Jing Lu, Tao<br>Liu, Baisheng Li, Shuijiang Mei, Feng<br>Ruan, Lifeng Lin, Changwen Ke, Hao-<br>jie Zhong, Yingtao Zhang, Lirong<br>Zou, Xuguang Chen, Qi Zhu, Jian-<br>peng Xiao, Jianxiang Geng, Zhe Liu,<br>Jianxiang Hu, Weilin Zeng, Xing<br>Li, Yuhuang Liao, Xiujuan Tang,<br>Songjian Xiao, Ying Wang, Yingchao<br>Song, Xue Zhuang, Liyun Liang,<br>Guanhao He, Huihong Deng, Tie Song,<br>Jianfeng He, Wenjun Ma |

**Table 21.**
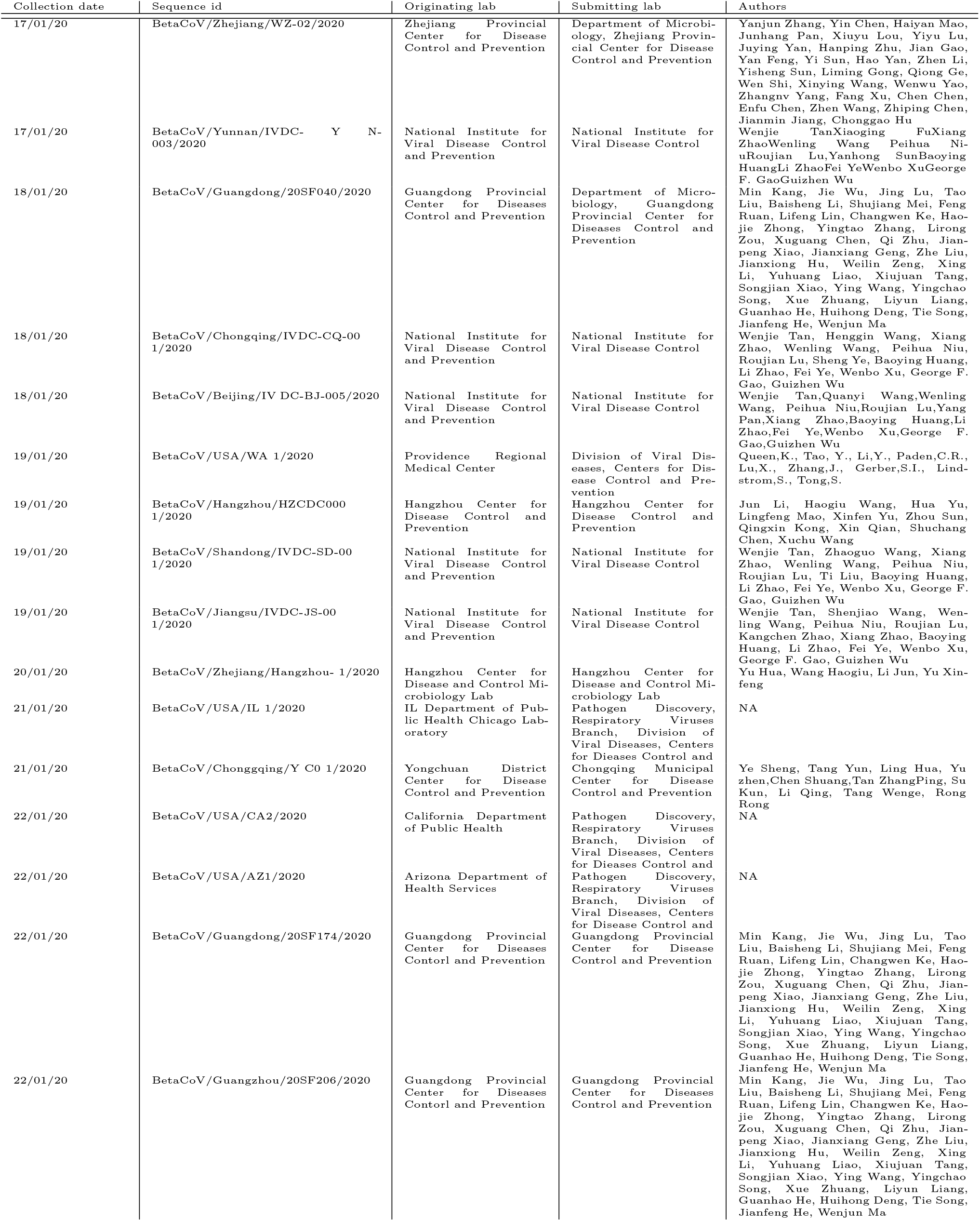
Nucleotide sequences used in this study that were obtained from the GISAID EpiFlu Database™(contd.).

**Table 22.**
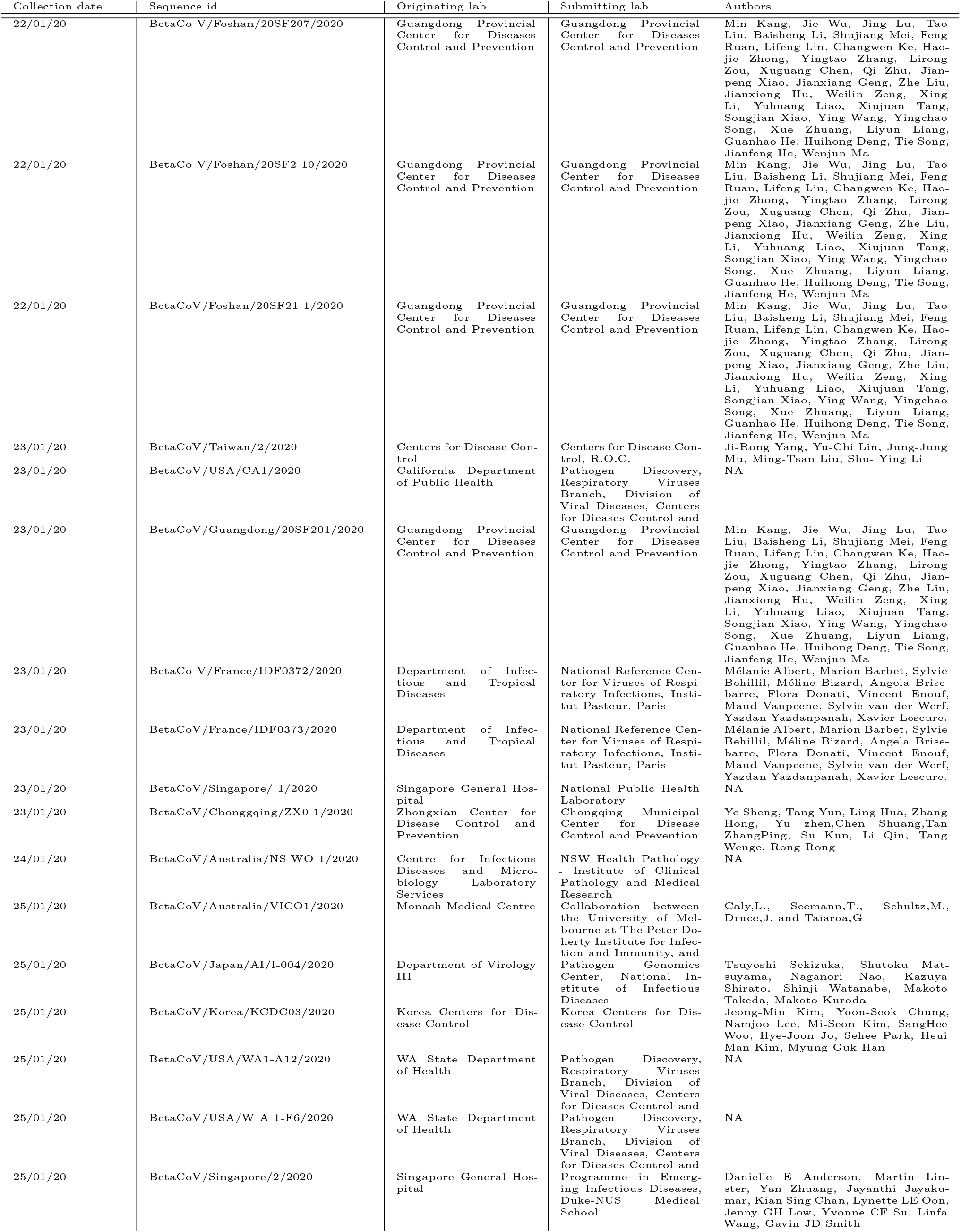
Nucleotide sequences used in this study that were obtained from the GISAID EpiFlu Database™(contd.).

**Table 23.** Nucleotide sequences used in this study that were obtained from the GISAID EpiFlu Database™(contd.).

| Collection date | Sequence id | Originating lab | Submitting lab | Authors |
| --- | --- | --- | --- | --- |
| 28/01/20 | BetaCoV/Germany/BavPat 1/2020 | Charité Universitätsmedizin Berlin | Charité Universitätsmedizin Berlin, Institute of Virology | Victor M Corman, Julia Schneider, Talitha Veith, Barbara Miihlemann, Markus Antwerpen, Christian Drosten, Roman Wolfel |
| 28/01/20 | BetaCoV/Australia/QLD001/2020 | Pathology Queensland | Public Health Virology Laboratory | Ben Huang, Alyssa Pyke, Amanda De Jong, Andrew Van Den Hurk, Carmel Taylor, David Warrillow, Doris Genge, Elisabeth Gamez, Glen Hewitson, Ian Maxwell Mackay, Inga Sultana, Jamie McMahon, Jean Barcelon, Judy Northill, Mitchell Finger, Natalie Simpson, Neelima Nair, Peter Burtonclay, Peter Moore, Sarah Wheatley, Sean Moody, Sonja Hall-Mendelin, Timothy Gardam, and Frederick Moore. |
| 29/01/20 | BetaCoV/England/01/2020 | Respiratory Virus Unit | Respiratory Virus Unit, Microbiology Services Colindale, Public Health England | Monica Galiano, Shahjahan Miah, Richard Myers, Angie Lackenby, Omolola Akinbam1, Tiina Talts, Leena Bhaw, Kirstin Edwards, Jonathan Hubb, Joanna Ellis, Maria Zambon |
| 29/01/20 | BetaCoV/England/02/2020 | Respiratory Virus Unit | Respiratory Virus Unit, Microbiology Services Colindale, Public Health England | Monica Galiano, Shahjahan Miah, Richard Myers, Angie Lackenby, Omolola Akinbam1, Tiina Talts, Leena Bhaw, Kirstin Edwards, Jonathan Hubb, Joanna Ellis, Maria Zambon |
| 29/01/20 | BetaCoV/Finland/1/2020 | Lapland Central Hospital | Department of Virology, University of Helsinki and Helsinki University Hospital, Helsinki, Finland | Teemu Smura, Suvi Kuivanen, Hanni-mari Kallio-Kokko, Olli Vapalahti |
| 29/01/20 | BetaCoV/USA/CA3/2020 | California Department of Health | Pathogen Discovery, Respiratory Viruses Branch, Division of Viral Diseases, Centers for Disease Control and | NA |
| 29/01/20 | BetaCoV/USA/CA4/2020 | California Department of Health | Pathogen Discovery, Respiratory Viruses Branch, Division of Viral Diseases, Centers for Disease Control and | NA |
| 29/01/20 | BetaCoV/USA/CA5/2020 | California Department of Health | Pathogen Discovery, Respiratory Viruses Branch, Division of Viral Diseases, Centers for Disease Control and | NA |
| 29/01/20 | BetaCoV/France/IDF05 15/2020 | Department of Infectious and Tropical Diseases | National Reference Center for Viruses of Respiratory Infections, Institut Pasteur, Paris | Mélanie Albert, Marion Barbet, Sylvie Behillil, Méline Bizard, Angela Brisebarre, Flora Donati, Vincent Enouf, Maud Vanpeene, Sylvie van der Werf, Yazdan Yazdanpanah, Xavier Lescure |
| 29/01/20 | BetaCoV/France/IDF0626/2020 | Sorbonne Université | National Reference Center for Viruses of Respiratory Infections, Institut Pasteur, Paris | Mélanie Albert, Marion Barbet, Sylvie Behillil, Méline Bizard, Angela Brisebarre, Flora Donati, Vincent Enouf, Maud Vanpeene, Sylvie van der Werf, Sonia Burrel, Anne-Genevieve Marcelin, Vincent Calvez, David Boutolleau, Elise Klément, Valérie Pourcher, Eric Caumes. |
| 30/01/20 | BetaCoV/Australia/QLD02/2020 | Pathology Queensland | Public Health Virology Laboratory | Ben Huang, Alyssa Pyke, Amanda De Jong, Andrew Van Den Hurk, Carmel Taylor, David Warrillow, Doris Genge, Elisabeth Gamez, Glen Hewitson, Ian Maxwell Mackay, Inga Sultana, Jamie McMahon, Jean Barcelon, Judy Northill, Mitchell Finger, Natalie Simpson, Neelima Nair, Peter Burtonclay, Peter Moore, Sarah Wheatley, Sean Moody, Sonja Hall-Mendelin, Timothy Gardam, and Frederick Moore. |
| 31/01/20 | BetaCoV/Taiwan/NTU01/2020 | Department of Laboratory Medicine | Microbial Genomics Core Lab, National Taiwan University Centers of Genomic and Precision Medicine | Shiou-Hwei Yeh, You-Yu Lin, Ya-Yun Lai, Chiao-Ling Li, Shan-Chwen Chang, Pei-Jer Chen, Sui-Yuan Chang |
| 1/02/20 | BetaCoV/Singapore/3/2020 | National Centre for Infectious Diseases | Programme in Emerging Infectious Diseases, Duke-NUS Medical School | Danielle E Anderson, Martin Linster, Yan Zhuang, Jayanthi Jayakumar, David CB Lye, Yee Sin Leo, Barnaby E Young, Yvonne CF Su, Linfa Wang, Gavin JD Smith |
| 3/02/20 | BetaCoV/Belgium/GHB-0302 1/2020 | KU Leuven | KU Leuven, Clinical and Epidemiological Virology | Bert Vanmechelen, Elke Wollants, Annabel Rector, Els Keyaerts, Lies Laenen, Marc Van Ranst, and Piet Maes |

## Footnotes

1 https://www.gisaid.org/

2 https://github.com/vuw-c2lab/transcendental-information-cascades

3 https://www.gisaid.org/

